# RBMX functional retrocopy safeguards brain development

**DOI:** 10.1101/2025.10.17.25337589

**Authors:** Pierre Tilliole, Carolin Mattausch, Peggy Tilly, Elsa Leitão, Lucile Boutaud, Daphné Lehalle, Isabelle An, Emanuela Argilli, Sharon Aufox, Bert Callewaert, Perrine Charles, Jessica K. Cinkornpumin, Thomas Courtin, Marco Dalla Vecchia, Erica E. Davis, Boyan Ivanov Dimitrov, William Dobyns, Ekaterina Epifanova, Erwan Grandgirard, Matthieu Jung, Sarah Jurgensmeyer Langas, Sabine Kaya, Boris Keren, Tahir N. Khan, Elodie Lejeune, Mingfeng Li, Yannick Marie, Bastien Morlet, Caroline Nava, William A. Pastor, Damien Plassard, Carlos E. Prada, Agnès Rastetter, Noémie Schwaller, Nenad Sestan, Elliott Sherr, Suzanna L. Temple, Jude-Felix Tenywa, Sylvia Tielens, Arie van Haeringen, Helen Whitley, Laurent Nguyen, Laura Steenpaß, Muriel Rhinn, Stephan C. Collins, Delphine Héron, Valerie Cormier-Daire, Tania Attie-Bitach, Binnaz Yalcin, Christel Depienne, Juliette D. Godin

**Author notes:** These authors contributed equally to this work (Christel Depienne, Juliette D. Godin).

## Abstract

Retrotransposition has generated thousands of intronless gene copies in mammalian genomes, yet their contribution to brain development and evolution remains largely unexplored. Here we uncover a critical role for RBMX retrocopy in shaping neurodevelopment and modulating disease. Pathogenic variants in *RBMX*, an X-linked splicing regulator, cause intellectual disability, microcephaly, and cortical malformations. Through integrated human genetic, cellular, and mouse model studies, we show that *RBMX* pathogenic variants disrupt cortical development through both loss- and gain-of-function mechanisms. Surprisingly, despite severe phenotypes in humans, *Rbmx*-deficient mice display only mild cortical abnormalities, a discrepancy likely due to compensation by *Rbmxl1*, a retrocopy that arose independently in mice and humans. We demonstrate that RBMX and RBMXL1 share protein and RNA partners and act redundantly in brain development, with RBMXL1 buffering the impact of *RBMX* deficiency. These findings establish retrocopies as functional paralogs safeguarding neurodevelopment, and suggest that functional RBMX retrocopy could contribute to the robustness and evolutionary diversification of mammalian brain.

## INTRODUCTION

*RBMX* is a gene located on chromosome X in mammals that plays various roles in regulating gene transcription and alternative splicing.^1^ This gene encodes several isoforms resulting from alternative splicing. The main and longer protein isoform, also known as heterogeneous nuclear ribonucleoprotein (hnRNP) G, is a component of the spliceosome machinery.^2^ This 391 amino-acid protein contains two different RNA-binding domains at its N-terminus (RNA recognition motif, RRM) and C-terminus.^1,2^ HnRNP G/RBMX specifically recognises the CC(A/C) consensus motif and regulates the inclusion or exclusion of alternatively spliced exons where this motif occurs in close proximity, such as exon 7 of *SMN2*.^2^ RBMX also represses the selection of cryptic splice sites, especially within long exons, as evidenced for exon 9 of *ATRX*.^3^ In more distant species like zebrafish and *Xenopus*, the *rbmx* orthologue is located on autosomes^4^ and its knockdown during embryogenesis leads to a range of developmental abnormalities in the brain, eyes, and muscles, underscoring the critical role of *RBMX* in the development of several organs, particularly the brain.^4,5^

Remarkably, *RBMX* has multiple genomic paralogues in mammals. These paralogues include duplicated genes with similar intron/exon structures on the Y chromosome, which have evolved from an ancestral X/Y pair of chromosomes.^1,6^ In humans, six functional, almost identical copies and more than 20 pseudogenes exist on the long arm of chromosome Y. Unlike *RBMX,* which is expressed ubiquitously, *RBMY* genes are expressed specifically in germ cells, where they play a key role during spermatogenesis, and their deletion leads to male infertility.^1,7,8^ In addition, *RBMX* has been retrocopied several times during evolution, with at least nine intronless copies in the human genome. Several ancient retrocopies are non-functional (*RBMXP1-4*), while others remain functional with specific expression patterns (e.g., testis-specific *RBMXL2* and *RBMXL3*) or functions (e.g., *RBMX2*).^4^ More recent retrocopies have been described in human (*RBMXL1*) and mouse (*Rbmxl1*).^6,9^ Their high (>95%) protein sequence identity suggests that *RBMXL1* retrocopy is functional and could compensate for the loss of *RBMX* function.

Due to the presence of copies and retrocopies in human genomes, analysing *RBMX* using short-read sequencing is challenging and has likely limited the identification of pathogenic variants. To date, only two *RBMX* variants have been reported in two large families with X-linked intellectual disability (ID) syndromes. Shashi syndrome (MIM #300238) is characterised clinically by moderate ID, obesity, and dysmorphic facial features, and was associated with a hemizygous 23 bp deletion that leads to a frameshift in the final exon.^10^ Gustavson syndrome (MIM #309555)^11^ is characterised by more severe symptoms including profound ID, visual loss, hearing impairment, spasticity, seizures and early death, and was associated with the in-frame deletion of proline 162.^12^ These variants possibly have different functional consequences: the Shashi variant was suggested to result in *RBMX* loss-of-function^13^, while deletion of Pro162 was reported to alter RBMX interaction with SH3 binding proteins.^12^ However, as only two families with *RBMX* variants associated with distinct phenotypes have been reported, further evidence is needed to confirm the gene’s association with any of these syndromes and determine the associated pathogenic mechanisms.

In this study, we provide definitive evidence that pathogenic *RBMX* variants cause a spectrum of neurodevelopmental disorders in humans, driven by opposing molecular mechanisms and partially buffered by its autosomal retrocopy, *RBMXL1*.

## RESULTS

### *RBMX* variants cause severe X-linked neurodevelopmental disorders in hemizygous males

Using exome sequencing, GeneMatcher, and international collaborations, we assembled a cohort of 11 individuals from nine unrelated families harboring pathogenic or likely pathogenic *RBMX* variants (**Fig. 1a,b**). Four of the eight distinct variants introduced premature termination codons in the last exon of the MANE isoform (NM_002139.3). The c.1063dup (p.Arg355Lysfs*8) variant was found in the hemizygous state in two brothers with microcephaly, agenesis of the corpus callosum, microphthalmia, spasticity, and profound ID (Family 7). The same variant also occurred *de novo* in an unrelated sporadic case (Family 8). The c.1057_1058del (p.Met353Glyfs*9) variant was found hemizygous in a male patient with a thin corpus callosum, developmental delay, and behavioral disturbances (Family 6). Notably, c.1057_1058del (p.Met353Glyfs*9), c.1063dup. (p.Arg355Lysfs*8), and c.1037_1059del (p.Glu346Glyfs*9), previously reported in Shashi syndrome,^10,14^ all cause nearly similar frameshifts that add a conserved stretch of seven residues (KGVPSST; **Fig. 1c**). Two variants were nonsense: c.1141C>T (p.Arg381*), segregating in five affected individuals from an X-linked family who exhibited ID, microcephaly, microphthalmia, arthrogryposis or spasticity, and seizures (Family 9); the c.1033C>T (p.Gln345*) variant was identified in a patient with microcephaly, microphthalmia, dysplastic corpus callosum, brain malformation, and abnormal genitalia (Family 5, **Supplementary Fig. 1a**). The remaining four variants included the in-frame deletion of proline 162 (c.484_486del, p.Pro162del; Family 4, **Supplementary Fig. 1b**) already described in Gustavson syndrome^12^, a novel in-frame deletion (c.247_249del, p.Gln83del; Family 3), and two missense variants (c.37G>C, p.Gly13Arg, Family 1; and c.166T>C, p.Phe56Leu; Family 2; **Supplementary Fig. 1c,d**). These three later variants occurred *de novo* in isolated cases and affect highly conserved residues of the N-terminal RRM domain (**Fig. 1b**). p.Gly13Arg was found in a patient with epileptic encephalopathy; p.Gln83del in a patient with a thin corpus callosum, developmental delay, short stature, scoliosis, and facial dysmorphism; and p.Phe56Leu in a patient with microcephaly, growth delay, hearing impairment, skeletal anomalies, and underdeveloped genitalia. All variants are absent from gnomAD v.4.1, and predicted pathogenic by various prediction scores, including CADD PHRED scores (**Supplementary Table 2**).

**Fig. 1.**
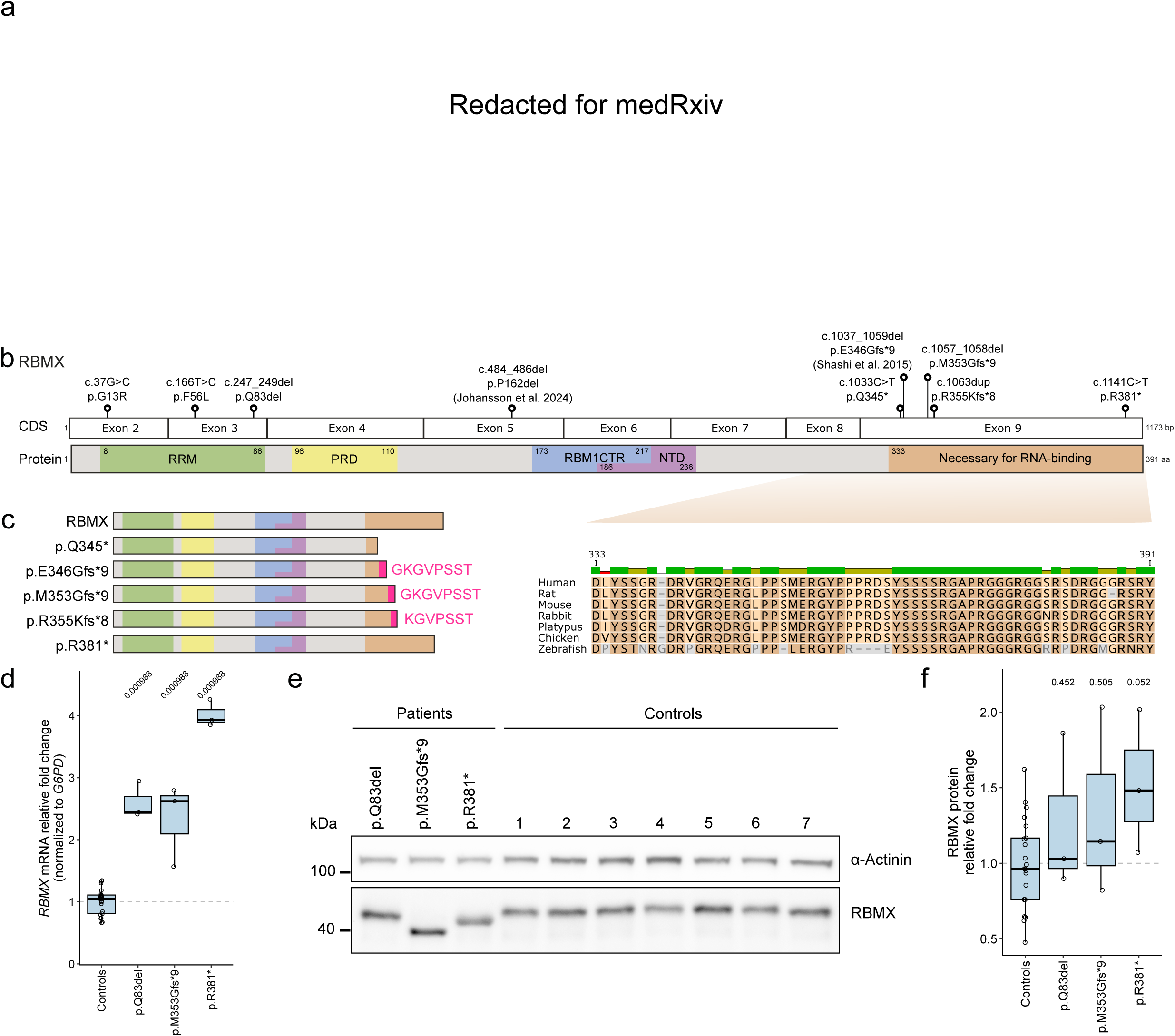
Spectrum of *RBMX* variants associated with neurodevelopmental disorders. **a,** retracted for MedRxiv **b**, Schematic representation of the variants on the *RBMX* mRNA (above, boxes indicate exons) and protein (below; colored boxes indicate the RBMX functional domains). A high degree of conservation of the RNA-binding domain is observed, extending up to zebrafish. **c**, Schematic representation of the impact of truncating variants (frameshift and nonsense mutations) on the RBMX protein. All frameshift variants result in the same reading frame shift, leading to the addition of almost identical stretches of aberrant amino acids. **d,** *RBMX* expression in patient fibroblasts versus controls measured by RT-qPCR. *RBMX* expression was normalized to *G6PD* and to the mean of controls per experiment (*n*=3 / three technical replicates each). Mann-Whitney U test was used to determine *P*-values indicated above comparisons. **e**,**f**, Immunoblot (**e**) and quantification (**f**) of RBMX and RBMXL1 in patient and control fibroblasts. **e,** Representative Western blot of RBMX/RBMXL1 in fibroblasts from patients with indicated RBMX variants (first three lanes), age-matched controls (controls 1-4), and adult controls (controls 5-7). α-Actinin served as a loading control; molecular masses in kilodaltons (kDa). **f**, Quantification of band intensities shown as RBMX to α-Actinin ratios normalized to the mean of controls (*n*=3). Comparisons were performed with Mann–Whitney U test. *P*-values are shown above for comparisons.

For three patients (p.Gln83del, p.Met353Glyfs*9, and p.Arg381*), fibroblast samples were available. *RBMX* expression was assessed by quantitative RT-PCR and Western blotting, revealing a 2.5–4-fold increase in mRNA levels (**Fig. 1d**, **Supplementary Fig. 2a,b**), without a comparable increase in protein levels relative to sex-matched control fibroblasts (**Fig. 1e,f**). Both Met353Glyfs*9 and p.Arg381* produced truncated proteins lacking the conserved C-terminal region of the RNA-binding domain (**Fig. 1e**). Like wild-type (wt) RBMX, all mutant proteins are located within the nucleus (**Supplementary Fig. 2c**). Altogether, these findings confirm that pathogenic *RBMX* variants are responsible for a spectrum of neurodevelopmental disorders, most commonly characterized by intellectual disability, microcephaly, and corpus callosum anomalies. Variants fall into two main categories: truncating changes in the last exon resulting in C-terminally truncated proteins, and missense or in-frame variants in the N-terminal region.

### RBMX retrocopy events in mouse and human are independent

*RBMX-*related disorders exhibit significant phenotypic variability both within and between families. Given the presence of multiple *RBMX* copies and retrocopies, we hypothesized that a closely related copy may compensate for *RBMX* function, potentially explaining the clinical variability. To investigate the evolutionary history of *RBMX* and its retrocopies, we retrieved the closest *RBMX* orthologues and paralogues (**methods**) and reconstructed the phylogenetic relationships among *RBMX*-related genes across vertebrates (**Fig. 2a**). This analysis confirmed that *Rbmxl2* diverged early in vertebrate evolution, prior to the split between fish and tetrapods. Intronless *RBMX* retrocopies are found exclusively in placental mammals, and several retrotransposition events occurred independently throughout evolution. The human closest retrocopy, *RBMXL1,* encodes a protein 96% identical to RBMX (16 amino acid differences) and is embedded in intron 2 of *KYAT3* (kynurenine–oxoglutarate transaminase 3) on chromosome 1p22.2, sharing 5’ non-coding and regulatory regions with this gene (**Fig. 2b**). *RBMXL1* is only present in humans, apes, and Old World monkeys, confirming that this event is evolutionary recent.^6,9^

**Fig. 2.**
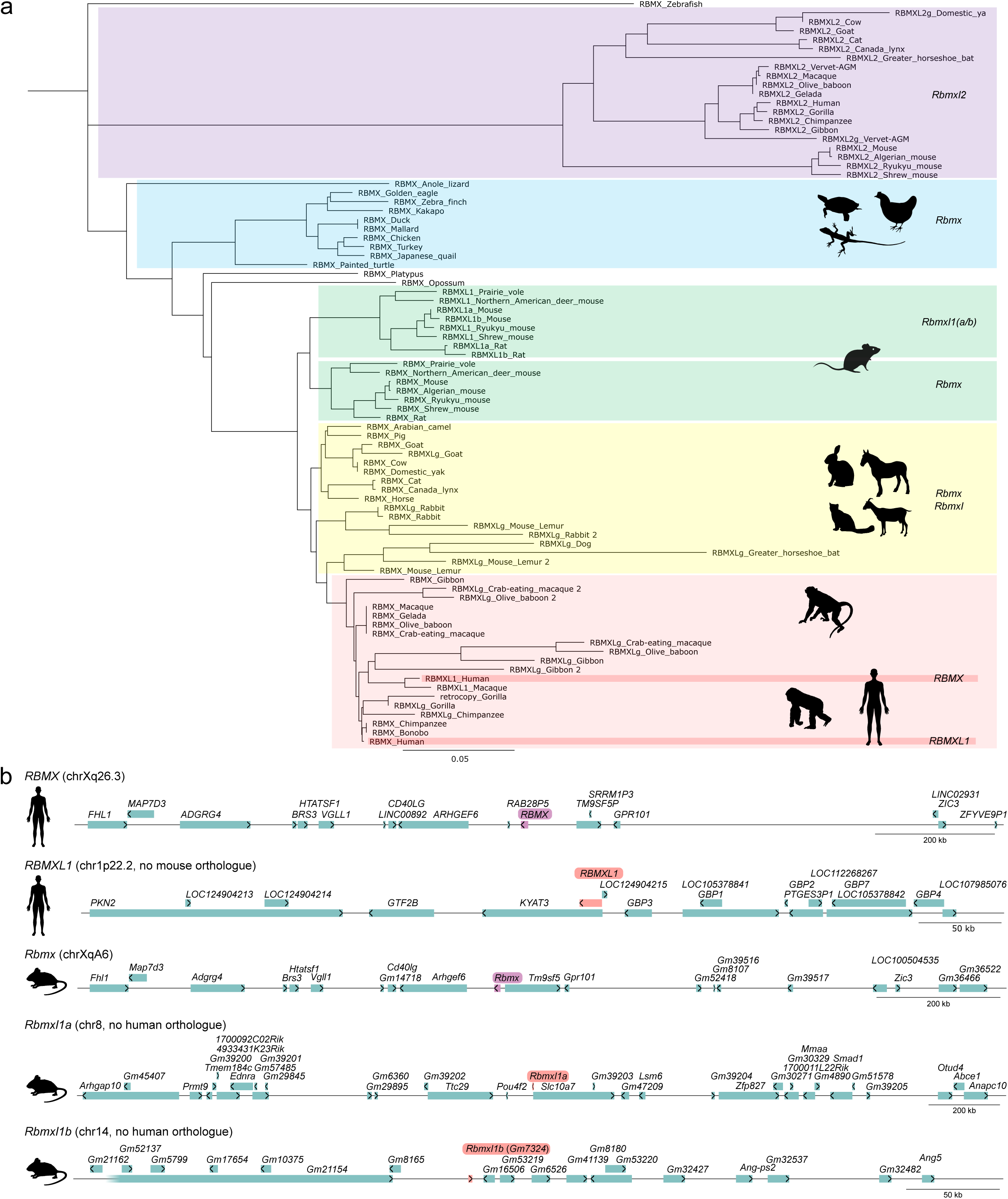
Human *RBMXL1* and mouse *Rbmxl1a/b* retrocopies originate from independent retrotransposition events. **a**, Phylogenetic tree showing *RBMX* orthologues and closely related retrocopies across selected species. Scale bar represents the number of substitutions per site. **b**, Genomic locations of *RBMX* and retrocopies in human (top) and mouse (bottom) genomes. Species-specific insertion sites support their independent origins.

In contrast, the mouse genome harbors two *Rbmxl1* retrocopies, each encoding proteins 98% identical to mouse *Rbmx* (**Fig. 2a; Supplementary Fig. 3a**). Both retrocopies are located in genomic contexts different than human *RBMXL1*: one, which we named *Rbmxl1a* - is located on chromosome 8, between *Pou4f2* and *Slc10a7* in a region syntenic to human chromosome 4. The other (*Gm7324,* renamed *Rbmxl1b*) is located on chromosome 14, between *Gm8165* and *Gm16506,* in a region lacking a syntenic counterpart in humans (**Fig. 2b**). Human *RBMXL1* and mouse *Rbmxl1a/Rbmxl1b* share a single missense change compared to *RBMX/Rbmx* while all other changes differ (**Supplementary Fig. 3b**), confirming that these retrocopies originated from independent retrotransposition events.

### *Rbmx* knockout mice show mild neuroanatomical defects reminiscent of those seen in humans

Through collaboration with the Sanger Institute Mouse Genetics Project^15^, we obtained samples from *Rbmx^tm2a^*mice. These were generated using the knockout-first allele method developed by the International Mouse Phenotyping Consortium (IMPC), in which exon 4 of *Rbmx* is floxed and a LacZ cassette inserted (**Fig. 3a**). The specific loss of *Rbmx*, but not *Rbmxl1*, in *Rbmx^tm2a^* mouse cortices was confirmed by RT-qPCR (**Supplementary Fig. 4a,b**). In 16-week-old mice, *Rbmx* is most highly expressed in the eye and testis, with only mild expression in the adult brain (**Supplementary Fig. 4c**). Despite this, *Rbmx^tm2a^* knockout male mice exhibited normal eye histology, with intact retinal layers (**Supplementary Fig. 4d**).

**Fig. 3.**
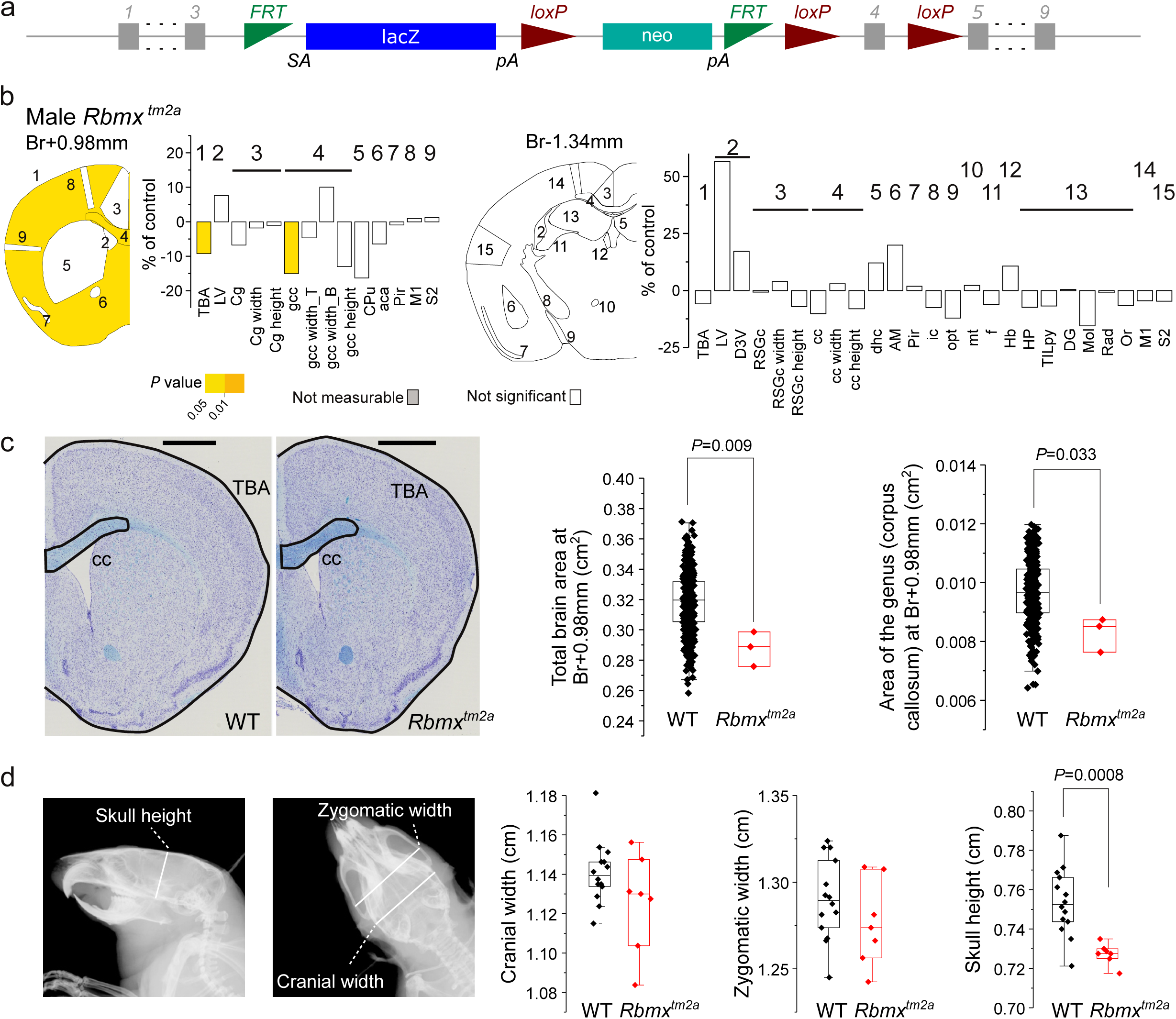
*Rbmx* deficiency is associated with mild neuroanatomical defects in the *Rbmx^tm2a^* mouse. **a,** Tm2a genetic construct showing the insertion of the LacZ and Neo cassettes upstream the critical exon 4 of *Rbmx*. **b**, Schematic representation of affected brain regions plotted in coronal planes according to *P*-values in three adult male *Rbmx^tm2a^*mice aged 16 weeks compared to matched wild-types mice (*n*=498). The left image represents parameters measured at Bregma +0.98 mm and the right image are parameters at Bregma −1.34 mm. Numbers indicate a group of studied brain regions. At Bregma +0.98 mm brain structures assessed were: the total brain area (TBA); the lateral ventricles (LV); the Cingulate cortex (Cg); the genu of the corpus callosum (gcc); the caudate putamen (CPu); the anterior commissure (aca); the piriform cortex (Pir); the primary motor cortex (M1); the secondary somatosensory cortex (S2). At Bregma −1.34 mm, a maximum of 18 brain structures were assessed: the total brain area (TBA); the lateral- (LV) and third ventricles (D3V); the retrosplenial granular cortex (RSGc); the corpus callosum (cc); the dorsal hippocampal commissure (dhc); the amygdala (AM); the piriform cortex (Pir); the internal capsule (ic); the optic tract (opt); the mammillothalamic tract (mt); the fimbria (f); the habenular nucleus (Hb); the hippocampus (HP); the total internal length of pyramidal cells (TILpy); the dentate gyrus (DG); the molecular layer of the hippocampus (Mol), the radiatum layer of the hippocampus (Rad); the oriens layer of the hippocampus (Or); the primary motor cortex (M1); the secondary somatosensory cortex (M2). The color code indicates the significance threshold, or white when not significant and grey when not computable. The full details of each neuroanatomical parameter and corresponding numbers are provided as **Supplementary Table 3**. Histograms represent percentage change relative to WT mice (set as 0) for each of the measured parameters. **c**, Representative Nissl-stained coronal brain sections from *Rbmx^tm2a^* mouse, showing the total brain area and the genu of the corpus callosum at Bregma (Br.) +0.98 mm in one hemisphere. Scale bar: 0.1 cm. Box plots of data are shown for the two significant parameters (black for *n*=498 WT and red for three mutants). The line in the middle of the box plots represents the median, the upper limit of the box corresponds to Q3, the lower limit to Q1, and the whiskers extend to 1.5× interquartile range. Data (means ± s.d.) was analyzed by two-tailed Student’s *t*-tests of equal variances. **d**, Cephalometric box plots for cranial and zygomatic width and skull height with overlay on X-ray showing each measurement (black for *n*=14 WT and red for seven *Rbmx^tm2a^* mutant mice). The differences were not statistically significant for cranial width (*P*=0.102) and zygomatic width (*P*=0,232). The line in the middle represents the median, the upper limit of the box corresponds to Q3, the lower limit to Q1, and the whiskers extend to 1.5× interquartile range. Data was analyzed by two-tailed Student’s *t*-tests of equal variances.

We next assessed brain morphology in 16-week-old *Rbmx^tm2a^*male mice using a standardized pipeline measuring 39 parameters across 24 brain regions at high resolution (**Supplementary Table 3**).^16^ Compared to wild-type controls, *Rbmx^tm2a^* mice exhibited only two neuroanatomical phenotypes including a 9.3% reduction in the total brain area (*P*-value=0.009) and a 15.1% decrease in the genu of the corpus callosum (*P*-value=0.033) in the first coronal section at Bregma +0.98 mm (**Fig. 3b,c**, **Supplementary Table 4**). The brain width reduction appeared more pronounced than height. Cephalometric X-ray analysis showed a significant 3.5% reduction in skull height (*P*-value=0.0008) and a 6% decrease in upper incisor height (*P*-value=5×10⁻⁵), while zygomatic and cranial widths were unchanged (**Fig. 3d, Supplementary Fig. 4e, Supplementary Tables 5-6**).

These results indicate that the brain anomalies in *Rbmx* knockout mice mirror the most frequent clinical brain features of *RBMX*-associated neurodevelopmental syndromes, namely microcephaly and corpus callosum anomalies. However, the abnormalities observed in mice are markedly milder than those in affected individuals, supporting the hypothesis that the two mouse retrocopies, *Rbmxl1a* and *Rbmxl1b*, provide greater compensation for the loss of *Rbmx* than the single human retrocopy *RBMXL1*.

### *RBMX* and *RBMXL1* exhibit molecular redundancy

We next sought to characterize the expression patterns of *RBMX* and *RBMXL1* in the developing brain. Immunostaining of coronal brain sections from a human fetus at gestational week (GW) 11 revealed widespread *RBMX* expression in the developing brain (**Fig. 4a**). Within the cortical wall, *RBMX/RBMXL1* were detected in both progenitors and neurons (**Fig. 4a**). However, due to the high protein sequence similarity between *RBMX* and its retrocopies, this antibody cross-reacts with both proteins, preventing its use for distinguishing individual protein expression.

**Fig. 4.**
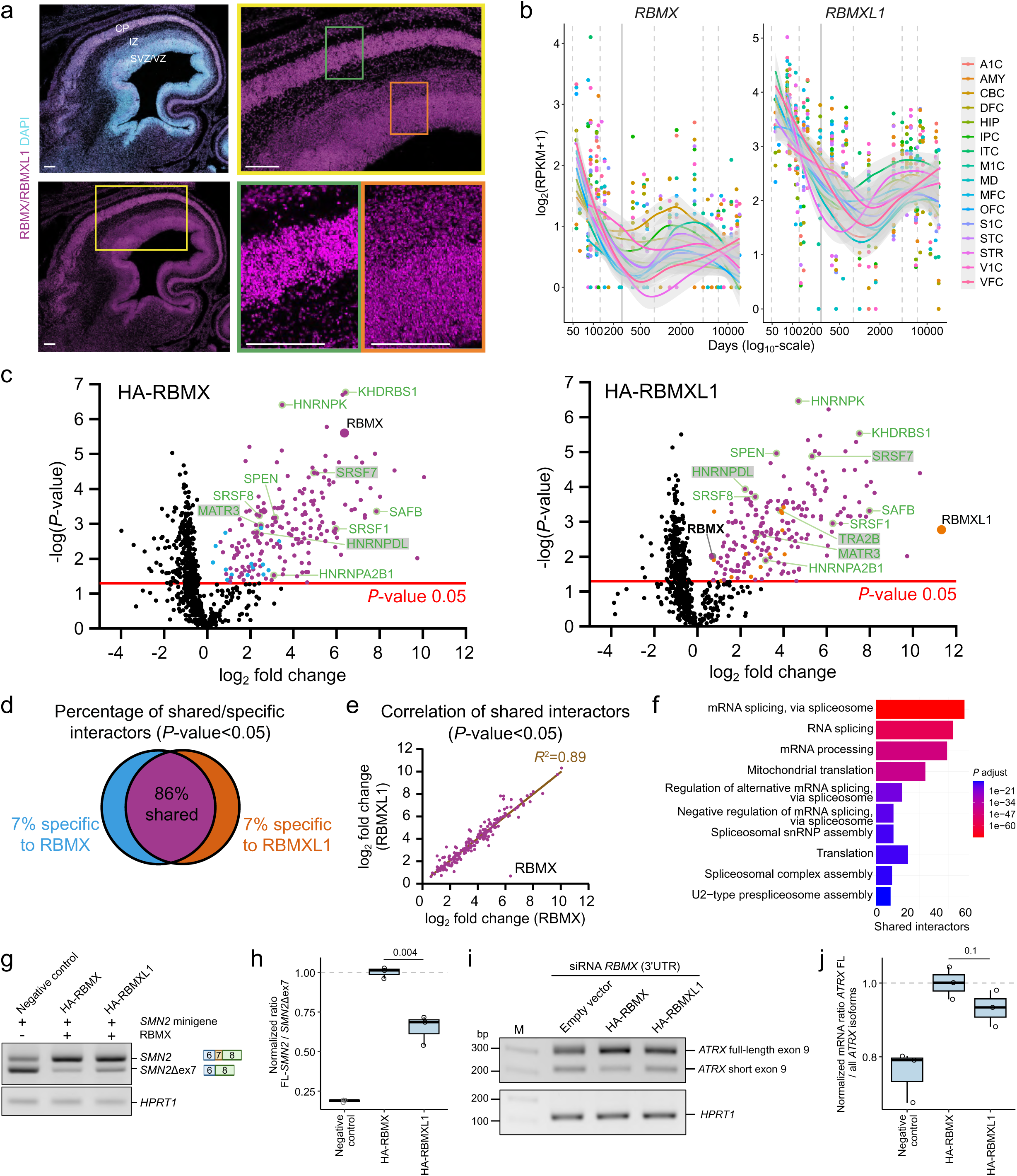
RBMX and its retrocopy RBMXL1 show overlapping expression and function in humans. **a**, Images of coronal human fetal brain section immunostained with RBMX/RBMXL1 antibody at GW11 (magenta). Nuclei are stained with DAPI (blue). Scale bars: 200 μm. VZ: ventricular zone, SVZ: subventricular zone, IZ: intermediate zone, CP: cortical plate. **b**, *RBMX* and *RBMXL1* expression (in log₂[RPMK + 1]; RPMK: Reads Per Kilobase per Million mapped reads) across multiple human brain regions, including A1C (primary auditory cortex), AMY (amygdala), CBC (cerebellar cortex), DFC (dorsolateral prefrontal cortex), HIP (hippocampus), IPC (inferior parietal cortex), ITC (inferior temporal cortex), M1C (primary motor cortex), MD (mediodorsal thalamus), MFC (medial prefrontal cortex), OFC (orbitofrontal cortex), S1C (primary somatosensory cortex), STC (superior temporal cortex), STR (striatum), VIC (visual cortex), and VFC (ventrolateral prefrontal cortex). Note that this representation does not allow direct comparison of expression levels between genes. **c**, Affinity purification-mass spectrometry results showing the proteins that bind significantly (*P-*value < 0.05, red line) to HA-RBMX (left, blue), HA-RBMXL1 (right, orange) or both (violet) in HEK293T cells. Shared interactors also identified in the yeast two-hybrid screen and validated by co-immunoprecipitation (**Supplementary Fig. 6b,c**) are highlighted in green and grey, respectively. Volcano plots show the negative log *P*-value of all proteins against their log_2_ fold change (log_2_FC) (HA-RBMX or HA-RBMXL1 versus empty HA vector). **d**, Venn diagram showing the percentage of shared and specific interactors among the protein partners identified in the mass spectrometry data (*P-*value < 0.05). **e**, Graph showing the correlation between the change in binding to RBMX and the change in binding to RBMXL1 for each shared interactor in log_2_ scales. **f**, Gene Ontology term analysis of shared interactors. Color represents the *P-*adjusted value. **g**, **h**, Detection (**g**) and quantification (**h**) of *SMN2* exon 7 inclusion in HEK293 cells co-expressing an *SMN2* minigene and an empty vector (negative control), HA-RBMX or HA-RBMXL1. **g**, Agarose gel showing PCR products for *SMN2* with exon 7 (*SMN2*) and *SMN2* without exon 7 (*SMN2Δex7*). *HPRT1* was used as a cDNA control. -: No HA-RBMX transfected, +: HA-tagged RBMX or RBMXL1 transfected. Schematic representations indicate *SMN2* exon 7 inclusion or skipping (exon 6: blue, exon 7: orange, exon 8: green). **h**, Boxplots of *SMN2* normalized to *SMN2Δex7* based on RT-qPCR data (*n*=3). Data are normalized to the mean of HA-RBMX samples. Statistical significance was assessed using Unpaired t-test. *P*-value is shown above for comparison (to HA-RBMX). **i**, **j**, Detection (**i**) and quantification (**i**) of *ATRX* exon 9 exitron inclusion in HEK293 cells after endogenous *RBMX* knockdown and rescue with an empty vector (negative control), HA-RBMX or HA-RBMXL1. (**i**) Agarose gel showing PCR products for *ATRX* with exitron inclusion (*ATRX* full-length exon 9) and with exitron exclusion (*ATRX* short exon 9). *HPRT1* is used as a cDNA control. (**j**) Boxplots of *ATRX* full-length exon 9 normalized to total *ATRX* exon 9 amounts based on RT-qPCR data (*n*=3). Data are normalized to the mean of HA-RBMX samples. Statistical significance was assessed using Unpaired t-test. *P*-value is shown above for comparison (to HA-RBMX).

We then leveraged untranslated region (UTR) sequences unique to human *RBMX* and *RBMXL1* to specifically evaluate their expression profiles in BrainSpan data.^17^ Both genes showed similar expression patterns in the brain, with high expression during prenatal stages, and lower levels in adulthood, albeit with considerable inter-individual variability (**Fig. 4b**). In parallel, single-cell transcriptomic datasets from the CZ CELLxGENE Discover platform^18^ (**methods**) confirmed aligned spatial and temporal expression across brain regions and developmental stages (**Supplementary Fig. 5a**). Furthermore, *RBMX* and *RBMXL1* display overlapping expression in progenitors, all subtypes of cortical neurons and glial cells in both developing and adult cortices, although *RBMXL1* levels were consistently lower (**Supplementary Fig. 5b**). We next assessed the co-expression of both transcripts in neurons by extracting *RBMX* and *RBMXL1* specific reads from a single cell transcriptomic dataset from human developing cortex at GW16.^19^ This analysis demonstrated that 73.4% of neurons co-expressed both *RBMX* and *RBMXL1* transcripts (**Supplementary Fig. 5c**). Finally, we quantified RBMX and RBMXL1 specific peptides by mass spectrometry after immunoprecipitation of endogenous proteins using an antibody recognizing both proteins. The analysis of the relative expression of RBMX and RBMXL1 further demonstrated higher RBMX abundance in human neural stem cells (hNSCs) and hNSCs-derived neurons (79-fold and 34-fold, respectively) (**Supplementary Fig. 5 d,e**). Altogether, these results confirm co-expression of *RBMX* and *RBMXL1* during early human brain development.

We next examined whether RBMX and RBMXL1 share protein interaction partners using two complementary approaches. First, we performed independent yeast two-hybrid (Y2H) screens using full-length human RBMX and RBMXL1 as baits against a human fetal brain cDNA library. In parallel, we overexpressed HA-tagged human RBMX or RBMXL1 in HEK293T cells and performed immunoprecipitation followed by mass spectrometry (**Fig. 4c**, **Supplementary Table 7)**. Y2H screens identified 29 protein partners for RBMX and 33 for RBMXL1, with 15 shared between them (**Supplementary Table 8**). Notably, RBMX interacts with both itself and RBMXL1 (**Fig. 4c**, **Supplementary Tables 7-8**). Co-expressing HA-RBMX and Flag-RBMXL1 in HEK293T cells and using anti-HA immunoprecipitation further confirmed that RBMX and RBMXL1 form heterodimers (**Supplementary Fig. 11a,b**). Among the top protein interactors identified in the mass spectrometry data (*P*-value < 0.05; **Fig. 4c**, **Supplementary Table 7**), 86 % were common to RBMX and RBMXL1 (**Fig. 4d**), with an almost perfect correlation in the ranking order of the interactors (**Fig. 4e**). Gene Ontology (GO) analysis of shared interactors confirmed enrichment for RNA splicing factors (**Fig. 4f**, **Supplementary Table 9**), which is consistent with the predominant nuclear localization of both proteins in hNSCs upon overexpression (**Supplementary Fig. 6a**). Notably, of the 140 splicing factors identified as shared interactors by mass spectrometry, 10 were also found to directly interact with RBMX and RBMXL1 in the Y2H screen (**Fig. 4c**, interactors in green, **Supplementary Table 8**). To validate these interactions, we performed anti-HA immunoprecipitation assays on extracts from HEK293T cells transfected with HA-RBMX or HA-RBMXL1 and confirmed comparable, RNA-independent binding of both proteins to MATR3, hnRNPDL, TRA2B and SRSF7, all involved in RNA splicing (**Supplementary Fig. 6b,c**).

We next investigated whether RBMXL1 regulates the same RNA targets as RBMX. We first assessed RBMXL1 binding to known RBMX targets, *SMN1/2*^20,21^, *ATRX*^3^, *HP1α*^22^ mRNA and *ASPM*, one the most highly enriched transcripts in a RBMX iCLIP-seq dataset^3^ (**Supplementary Table 10**). Using CLIP-qPCR with anti-HA antibodies in hNSC expressing either HA-RBMX or HA-RBMXL1, we demonstrated that RBMXL1 binds to similar targets as RBMX, with a comparable efficiency (**Supplementary Fig. 6d**).

RBMX is known to promote the inclusion of exon 7 in *SMN2* transcripts^20,21^. We adapted a *SMN2* minigene assay to test whether RBMXL1 is capable of carrying out the same splicing function. The results show that RBMXL1 also promotes inclusion of *SMN2* exon 7, though with reduced efficiency, approximately 65% that of RBMX (**Fig. 4g,h**, **Supplementary Fig. 7a-c**). RBMX also represses the exitron within *ATRX* exon 9 mRNA, promoting a longer isoform^3^ (**Supplementary Fig. 7d-f**). Using a long isoform-specific PCR, we demonstrated that both RBMX and RBMXL1 promote the longer isoform by 24 and 18%, respectively, and rescue the increased proportion of the short *ATRX* isoform caused by *RBMX* depletion in HEK293 cells (**Fig. 4i,j**, **Supplementary Fig. 7d-h**). Collectively, these findings suggest that RBMXL1, like RBMX, acts as a spliceosomal component with redundant roles in human splicing regulation.

### Functional compensation of RBMX and its retrocopy RBMXL1 in mouse corticogenesis

We next investigated *Rbmx* and *Rbmxl1a/b* (hereafter referred to as *Rbmxl1*) expression and their potential redundancy *in vivo* in mice. Immunostaining and Western blotting with an antibody recognizing both proteins revealed overlapping spatial and temporal expression patterns in the developing mouse brain, similar to humans (**Supplementary Fig. 8a-c**). Transcript-specific RT-qPCR confirmed stable expression of both genes during cortical development (**Supplementary Fig. 8d, Supplementary Fig. 4a**). Further gene-specific analysis of a published single-cell RNA-sequencing dataset of E13 mouse cortices^23^ further supported co-expression of *Rbmx* and *Rbmxl1* in mouse apical progenitors and neurons (**Supplementary Fig. 8e**).

To investigate the molecular redundancy between mouse *Rbmx* and *Rbmxl1*, we compared the global transcriptome of N2A neuroblastoma cells following targeted depletion of either *Rbmx* or *Rbmxl1* using pools of transcript-specific micro-RNAs (miRNAs) (**Supplementary Fig. 9a**). Differential gene expression analysis revealed an almost complete overlap between transcripts deregulated upon *Rbmxl1* knockdown and those affected by *Rbmx* depletion (**Supplementary Fig. 9b**), indicating that the mouse retrocopy, like its human counterpart, retains function similar to the ancestral gene.

We then used *in utero* electroporation (IUE) to individually or simultaneously knockdown *Rbmx* and *Rbmxl1* in wild-type mouse cortices at embryonic day (E) 13.5. miRNAs targeting *Rbmx*, *Rbmxl1* or both genes under the control of an ubiquitous CAG promoter (**Supplementary Fig. 9c**) were co-electroporated with a pCAG:H2B-eGFP vector to express a nuclear GFP in electroporated progenitors and their progeny. Two days post-IUE, cells depleted of either genes showed a distribution similar to control cells (IUE miRNA scramble), whereas combined *Rbmx/Rbmxl1* knockdown significantly increased the proportion of GFP+ cells delaminating in the intermediate zone (IZ) (+38% and +44% for miRNA #1 and miRNA #2, respectively; *P*-value<0.0001; **Fig. 5a,b**), without increased cell death. We then assessed the ability of human RBMX and RBMXL1 to restore neurogenesis defects induced by *Rbmx*/*Rbmxl1* concomitant knockdown. Co-electroporation of miRNA-insensitive human RBMX or RBMXL1 constructs fully rescued the neurogenesis defect induced by co-depletion of *Rbmx/Rbmxl1* by E15.5 (**Fig. 5c,d**, **Supplementary Fig. 9d,e**). Together, these data demonstrate that RBMX and RBMXL1 act redundantly to regulate corticogenesis in mammals.

**Fig. 5.**
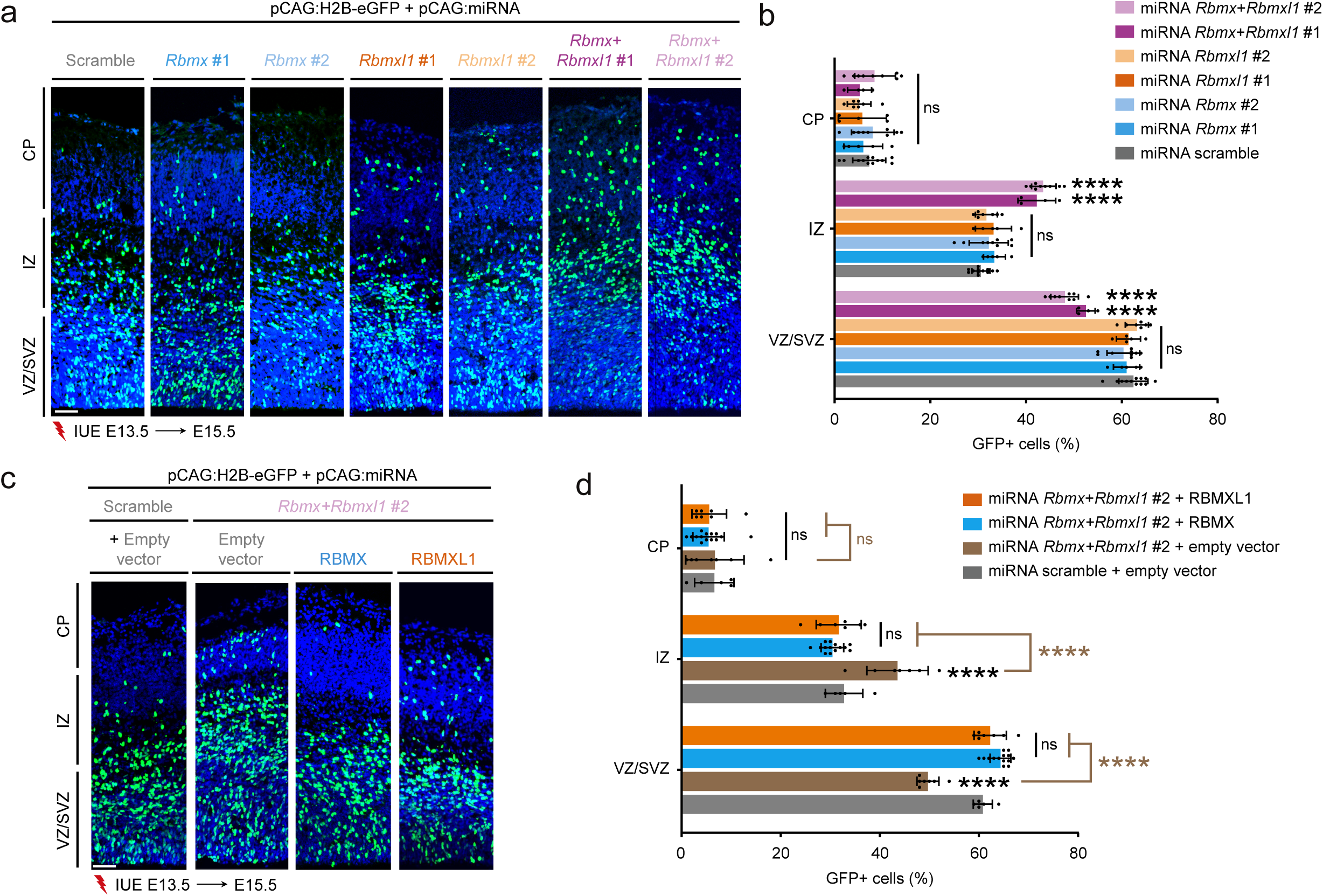
RBMX and its retrocopy RBMXL1 functionally compensate each other in neurodevelopment. **a**, Coronal sections of E15.5 mouse cortices, two days after *in utero* electroporation (IUE) with pCAG:miRNA scramble or pCAG:miRNAs targeting mouse *Rbmx* and *Rbmxl1* either individually (*Rbmx* #1 or #2; *Rbmxl1* #1 or #2) or simultaneously (*Rbmx+Rbmxl1* #1 or #2), together with pCAG:H2B-eGFP. GFP-positive electroporated cells are depicted in green. Nuclei are stained with DAPI (blue). Scale bar: 50 μm. **b**, Analysis of the percentage (means ± SEM) of electroporated cells in ventricular/subventricular zone (VZ/SVZ), intermediate zone (IZ), and cortical plate (CP) showing an accumulation of cells concomitantly depleted for *Rbmx* and *Rbmxl1* in IZ. Data were analyzed by Two-way ANOVA, with Tukey’s multiple comparisons test. Number of embryos analyzed: miRNA scramble, *n*=12; miRNA *Rbmx* #1, *n*=5; miRNA *Rbmx* #2, *n*=9; miRNA *Rbmxl1* #1, *n*=4; and miRNA *Rbmxl1* #2, *n*=7; miRNA *Rbmx+Rbmxl1* #1, *n*=4; miRNA *Rbmx+Rbmxl1* #2, *n*=9. ns: non-significant, \*\*\*\**P* < 0.0001. **c**, Coronal sections of E15.5 mouse cortices electroporated at E13.5 with pCAG:H2B-eGFP and pCAG:miRNA scramble or pCAG:miRNA *Rbmx+Rbmxl1* #2 together with empty vector or human WT RBMX or human WT RBMXL1. GFP-positive electroporated cells are depicted in green. Nuclei are stained with DAPI (blue). Scale bar: 50 μm. **d**, Analysis of the percentage (means ± SEM) of electroporated cells in ventricular/subventricular zone (VZ/SVZ), intermediate zone (IZ), and cortical plate (CP) showing the ability of the human WT RBMX and human WT RBMXL1 constructs to rescue the accumulation of cells depleted for mouse *Rbmx* and *Rbmxl1* in the IZ. Data were analysed by Two-way ANOVA, with Tukey’s multiple comparisons test. Number of embryos analyzed: miRNA scramble + empty vector, *n*=5; miRNA *Rbmx+Rbmxl1* #2 + empty vector, *n*=7; miRNA *Rbmx+Rbmxl1* #2 + RBMX, *n*=14; miRNA *Rbmx+Rbmxl1* #2 + RBMXL1, *n*=7. ns: non-significant, \*\*\*\**P* < 0.0001.

### RBMX variants perturb corticogenesis through two distinct mechanisms

We next aimed to elucidate the molecular mechanisms underlying the pathogenicity of *RBMX* variants. To evaluate their ability to rescue the neurogenesis defects induced by simultaneous co-depletion of *Rbmx* and *Rbmxl1*, we performed IUE of *Rbmx*/*Rbmxl1* miRNA #2 and pCAG:H2B-eGFP along with either human wild-type (WT) or mutant RBMX constructs in E13.5 embryos. We then analyzed the distribution of GFP-positive cells at E15.5. Under the conditions used in these complementation assays, neither WT nor mutant RBMX overexpression induced any phenotype in control conditions (scramble miRNA) (**Supplementary Fig. 10a,b**). Expression of WT-hRBMX and the p.Q83del variant fully restored the phenotype observed after *Rbmx*/*Rbmxl1* silencing (30% and 33% GFP+ cells in the IZ for WT and p.Q83del constructs, respectively, compared to 33% in control; *P*-value>0.05; **Fig 5c,d**, **Fig 6a,b**). In contrast none of the C-terminal truncated variants (p.Q345*, p.E346Gfs*9, p.R355Kfs*8 and p.R381*) nor p.P162del were able to rescue the defects (44%, 43%, 44%, 43%, 44% GFP+ cells in the IZ for the p.Q345*, p.E346Gfs*9, p.R355Kfs*8, p.R381* and p.P162del, respectively, compared to 44% in the knockdown condition; 0.0001<*P*-value<0.0011 (to control condition); **Fig. 6a,b**). These results suggest that C-terminal truncated proteins and p.P162del impair early neurogenic phase, at least partially, through a loss-of-function mechanism.

**Fig. 6.**
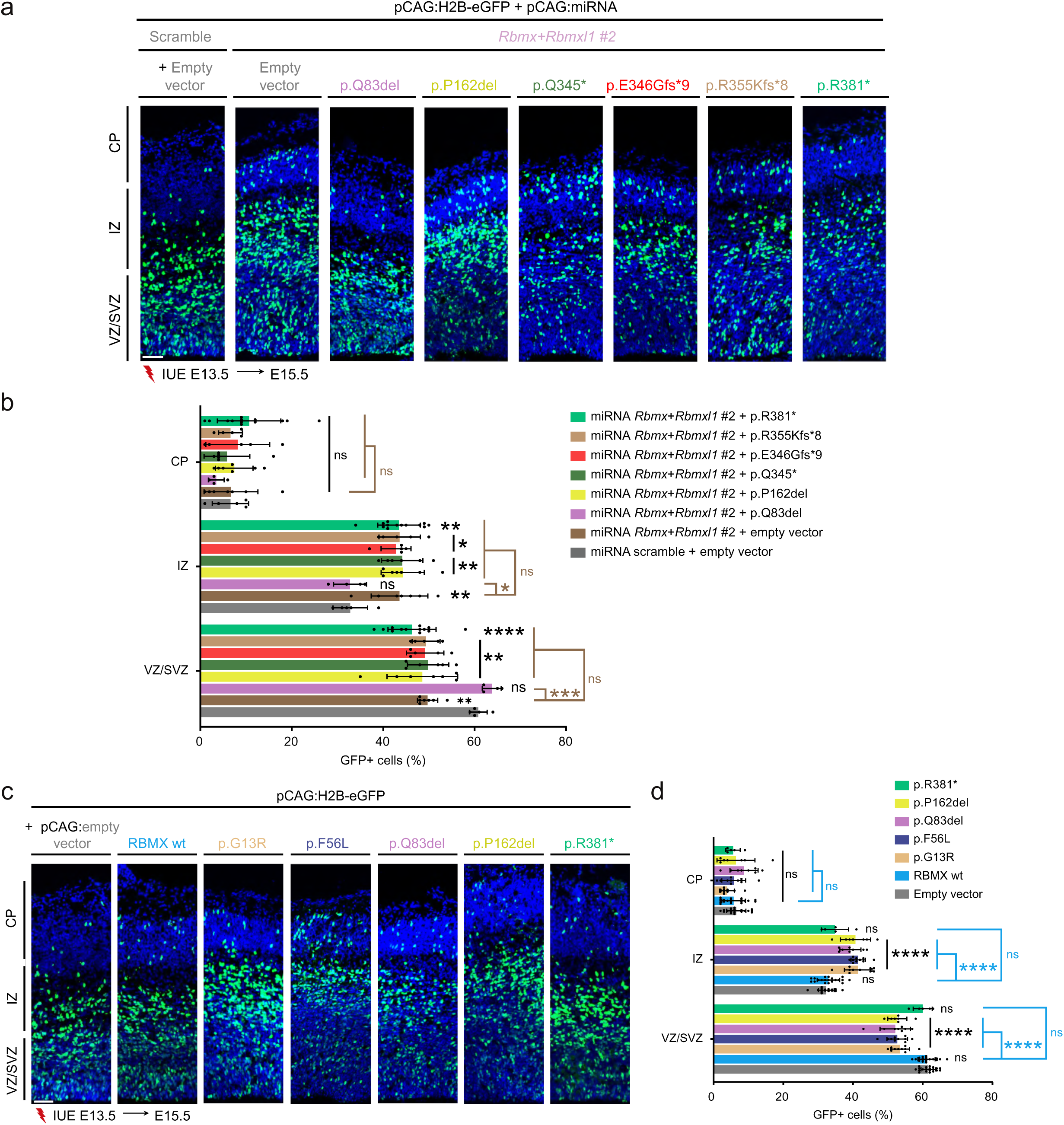
*RBMX* variants disrupt corticogenesis through both loss- and gain-of-function mechanisms. **a**, Coronal sections of E15.5 mouse cortices, two days after *in utero* electroporation (IUE) with pCAG:H2B-eGFP and pCAG:miRNA scramble or pCAG:miRNA *Rbmx+Rbmxl1* #2 together with empty vector or various variants as indicated. GFP-positive electroporated cells are depicted in green. Nuclei are stained with DAPI (blue). Scale bar: 50 μm. **b**, Analysis of the percentage (means ± SEM) of electroporated cells in ventricular/subventricular zone (VZ/SVZ), intermediate zone (IZ), and cortical plate (CP) showing the various abilities of the variants to rescue the accumulation of cells depleted for mouse *Rbmx* and *Rbmxl1* in the IZ. Data were analyzed by Two-way ANOVA, with Tukey’s multiple comparisons test. Number of embryos analyzed: miRNA scramble + empty vector, *n*=5; miRNA *Rbmx*+*Rbmxl1* #2 + empty vector, n=7; miRNA *Rbmx*+*Rbmxl1* #2 + p.Q83del, *n*=4; miRNA *Rbmx*+*Rbmxl1* #2 + p. P162del, *n*=7; miRNA *Rbmx*+*Rbmxl1* #2 + p.Q345*, *n*=6; miRNA *Rbmx*+*Rbmxl1* #2 + p.E346Gfs*9, *n*=5; miRNA *Rbmx*+*Rbmxl1* #2 + p.R355Kfs*8, *n*=5; miRNA *Rbmx*+*Rbmxl1* #2 + p.R381*, *n*=13. ns: non-significant, \**P* < 0.05, \*\**P* < 0.01, \*\*\**P* < 0.001, \*\*\*\**P* < 0.0001. **c**, Coronal sections of E15.5 mouse cortices, two days after *in utero* electroporation (IUE) with pCAG:H2B-eGFP and pCAG-empty vector or human WT RBMX, or various variants as indicated. GFP-positive electroporated cells are depicted in green. Nuclei are stained with DAPI (blue). Scale bar: 50 μm. **d**, Analysis of the percentage (means ± SEM) of electroporated cells in ventricular/subventricular zone (VZ/SVZ), intermediate zone (IZ), and cortical plate (CP) showing dominant effect of that variants located either in the N-terminal region (p.G13R, p.F56L, p.Q83del) or in the central region (p.162del) of *RBMX*. Data were analyzed by Two-way ANOVA, with Tukey’s multiple comparisons test. Number of embryos analyzed: empty vector, n=20; RBMX wt, *n*=2, p.G13R, *n*=10; p.F56L, *n*=10; p.Q83del, *n*=8; p. P162del, *n*=7; p.R381*, *n*=5. ns: non-significant, \*\*\*\**P* < 0.0001.

To further explore possible dominant effects, especially of variants located in the N-terminal RRM domain, we individually overexpressed RBMX WT, or p.G13R, p.F56L, p.Q83del, p.P162del, p.R381* variants in wild-type embryos at E13.5 using IUE. Except p.R381*, which behaved like WT (32%, 33%, 35%, GFP+ cells in the IZ for the control, WT and p.R381* constructs, respectively, *P*- value>0.05), all other variants significantly impaired GFP+ cell distribution, causing increased delamination into the IZ at E15.5 (42%, 42%, 39%, 41% GFP+ cells in the IZ for p.G13R, p.F56L, p.Q83del and p.P162del constructs, respectively; *P*-value<0.0001; **Fig. 6c,d**). Overall, these results reveal distinct pathogenic mechanisms for *RBMX* variants that depend on protein location. RRM domain variants likely act through a gain-of-function mechanism while C-terminal truncations cause a partial loss of-function. p.P162del exhibits a more complex mode of action with both loss-of-function and gain-of-function, possibly linked to cytoplasmic mislocalization of the corresponding protein (**Supplementary Fig. 10c**).

### RBMX mutants alter splicing of neurodevelopment-related genes

We next assessed the impact of RBMX variants on interaction and splicing *in vitro*. We first examined their ability to form heterodimers with RBMXL1. Y2H screens identified amino acids 133-292 as the minimal region required for autointeraction. Yet, anti-Flag immunoprecipitation assays in HEK293 cells co-transfected with WT or mutant HA-RBMX and Flag-RBMXL1 showed variants outside of this region also affected this interaction: N-terminal variants retain, and for some even enhance, the ability to interact with RBMXL1, whereas most C-terminal truncations severely impair heterodimer formation (**Supplementary Fig. 11a-d**). This indicates that regions spanning amino acids 133-292 and 355-381 are critical for auto-interaction and RBMXL1 binding. We then tested whether C-terminal truncation affects RBMX association with core spliceosome components. Anti-HA immunoprecipitation in HEK293T cells overexpressing WT or p.R381* HA-RBMX revealed that the truncated protein still associates with splicing factors (**Supplementary Fig. 11e,f)**, a result independently confirmed by a Y2H screen using human RBMX p.R381* as bait (**Supplementary Table 8**).

We then used a *SMN2* minigene assay to evaluate the splicing ability of RBMX variants. C-terminally truncated variants consistently reduced exon 7 inclusion (with values varying between 75-66% of WT), whereas p.G13R, p.F56L, and p.Q83del displayed variable splicing activity (45-77%). p.P162del again had an intermediate effect (56% inclusion) (**Fig. 7a,b**). RNA sequencing in fibroblasts from patients carrying p.Q83del (patient 3), p.M353Gfs*9 (patient 6) and p.R381* (patient 9-1), compared to sex and age-matched controls, revealed increased levels of the *ATRX* isoform with the exitron in the two patients with truncating variants but not in the patient with p.Q83del, suggesting a loss of exitron repression by C-terminal truncated proteins only (**Fig. 7c**). Exitron-specific PCR further confirmed a 2.6-fold increase in exitron-containing isoform in fibroblasts with p.R381* (**Fig. 7d-f**). The same effect was observed in genome-edited iPSCs with p.R355Kfs*8^13^ (**Fig. 7g-i**). Consistent with the loss-of-function effect of C-terminal truncated variants, depletion of both *RBMX* and *RBMXL1* in hNSCs similarly promoted the isoform with exitron (**Fig. 7j,k, Supplementary Fig. 11g,h**). In summary, our data demonstrate that distinct *RBMX* variants differentially affect splicing function in a domain- and target-specific manner, likely reflecting their diverse pathogenic mechanisms (**Fig. 6**). Finally, to directly test the role of RBMXL1 in modulating the phenotypic impact of RBMX variants, we performed *ATRX* exitron-specific PCR in fibroblasts from patient 9-1 (p.R381*) overexpressing either human RBMX or RBMXL1 (**Fig. 7l-m**). Strikingly, RBMXL1 overexpression rescues the *ATRX* splicing exitron, demonstrating that RBMXL1 can compensate for pathogenic RBMX variants and mitigate their molecular consequences.

**Fig. 7.**
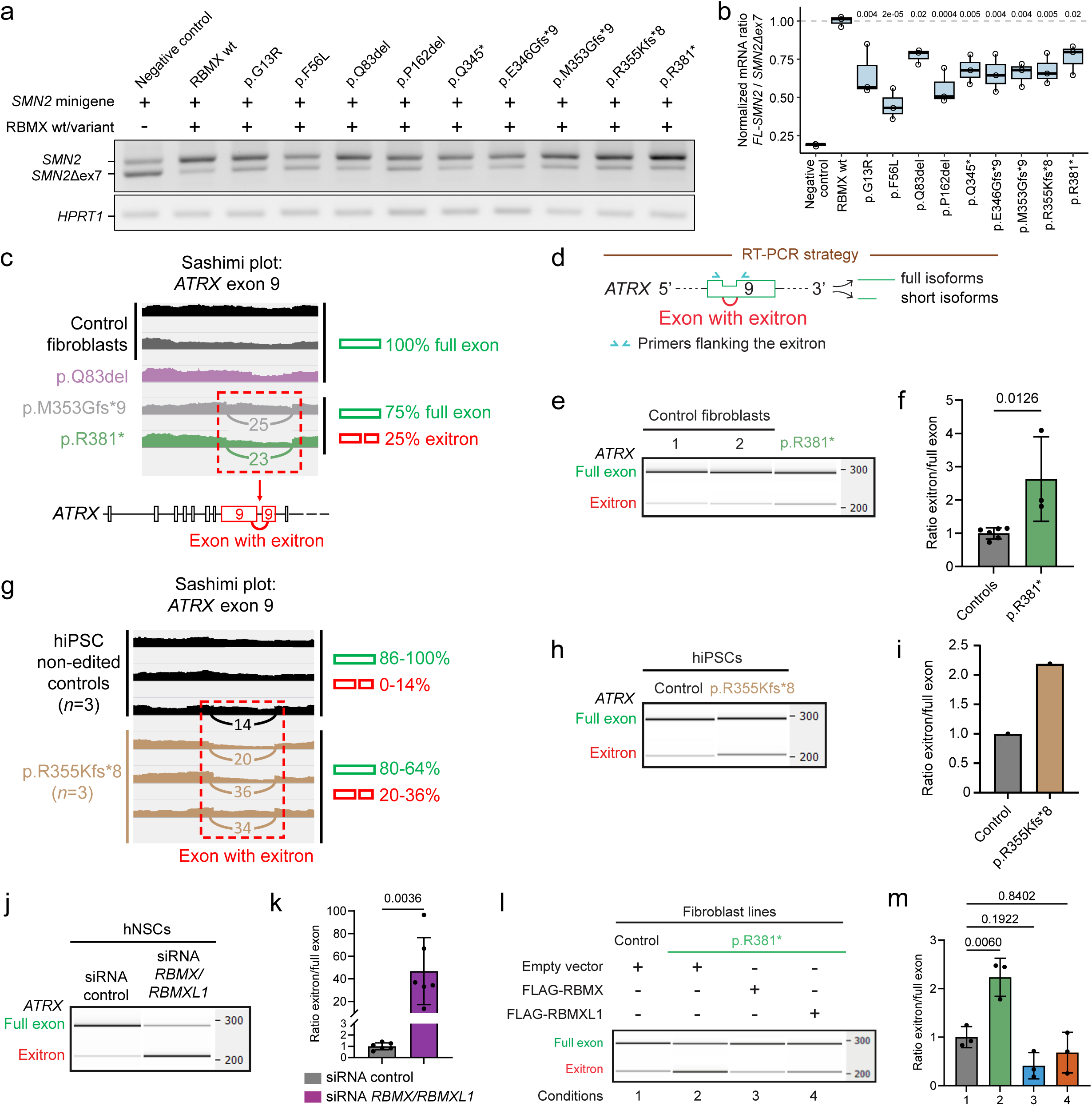
*RBMX* variants modulate splicing function in a domain- and target-specific manner. **a**, **b**, Detection (**a**) and quantification (**b**) of *SMN2* exon 7 inclusion in HEK293 cells co-expressing an *SMN2* minigene and an empty vector (negative control), HA-RBMX wildtype or variant as indicated. **a**, Agarose gel showing PCR products for *SMN2* with exon 7 (*SMN2*) and *SMN2* without exon 7 (*SMN2*Δex7). *HPRT1* is used as a cDNA control. -: No HA-RBMX transfected, +: HA-tagged RBMX wildtype or variant. **b**, Boxplots of *SMN2* normalized to *SMN2*Δex7 based on RT-qPCR data (*n*=3). Data are normalized to the mean of RBMX wt samples. Statistical significance was assessed using t-test and adjusted with Holm correction. Adjusted *P*-values are shown above for comparisons. **c**, Sashimi plot established from RNA-seq data of control fibroblasts and fibroblasts from patients 3 (p.Q83del), 6 (p.M353Gfs*9) and 9-1 (p.R381*) showing an increased proportion of *ATRX* exon 9 isoform with exitron in comparison to full-length *ATRX* exon 9 isoform upon C-terminal truncated RBMX variants expression. **d**, RT-PCR strategy using primers flanking the exitron in exon 9 to detect both short and full length *ATRX* isoforms. **e**, Representative gel electrophoresis of endogenous *ATRX* exon 9 analyzed by RT-PCR in fibroblasts from two controls and from patient 9-1 (p.R381*). **f**, Ratio between the short (exitron) and full-length (Full exon) *ATRX* mRNA isoforms expressed by the different fibroblast lines in (**e**). Data (means ± SEM) from 3 independent experiments were analyzed using Unpaired t-test. Data are normalized to the mean of control samples. *P*-value is indicated above for comparison. **g**, Sashimi plot established from available RNA-seq data^13^ of non-edited hiPSCs, *n*=3, and p.R355Kfs*8 edited hiPSCs, *n*=3, showing an increased proportion of *ATRX* exon 9 isoform with exitron in comparison to full-length *ATRX* exon 9 isoform in disease condition. **h**, Representative gel electrophoresis of endogenous *ATRX* exon 9 analyzed by RT-PCR in non-edited and p.R355Kfs*8 edited hiPSCs. **i**, Ratio between the short (exitron) and full-length (Full exon) *ATRX* mRNA isoforms expressed by the hiPSCs lines in (**h**) (*n*=1). **j**, Representative gel electrophoresis of endogenous *ATRX* exon 9 analyzed by RT-PCR in hNSCs transiently transfected with either a control siRNA or a siRNA targeting the ORF of *RBMX* and *RBMXL1*. **k**, Ratio between the short (exitron) and full-length (Full exon) *ATRX* mRNA isoforms expressed by the hiPSC lines in (**j**). Data (means ± SEM) from 6 independent experiments were analyzed using Unpaired t-test. Data are normalized to the mean of siRNA control samples. *P*-value is indicated above for comparison. **l,** Representative gel electrophoresis showing RT-PCR products of endogenous *ATRX* exon 9 from fibroblast lines (control and p.R381* (Patient 9-1)) transiently transfected with either an empty vector, FLAG-RBMX, or FLAG-RBMXL1 and FACS-sorted for mCherry-positive cells. **m**, Ratio between the short (exitron) and full-length (Full exon) *ATRX* mRNA isoforms expressed by the different fibroblast lines in (**l**). Data (means ± SEM) from 3 independent experiments were analyzed using one-way ANOVA and adjusted with Bonferroni correction. Data are normalized to the mean of control samples. Adjusted *P*-values are indicated above for comparison (to control condition).

## DISCUSSION

Genes with closely related pseudogenes or retrocopies, such as *RBMX*, pose significant challenges for clinical genetic analysis using short-read sequencing, likely contributing to the underdiagnosis of pathogenic variants in these regions. A decade after the first report linking a *RBMX* deletion to Shashi syndrome^10^, our study definitively establishes that *RBMX* variants cause a broad spectrum of neurodevelopmental disorders, ranging from moderate to profound intellectual disability. The most severe cases exhibit notable brain malformations, including mild to extreme microcephaly and corpus callosum anomalies. Additional but inconstant features include microphthalmia or microcornea, arthrogryposis or progressive spasticity, seizures, and genital defects. These findings underscore the critical role of *RBMX* in the development of the nervous system, eye, and testis, tissues where its expression is highest.

The spectrum of *RBMX* variants observed in patients, including in-frame or missense variants in the N-terminal region, as well as frameshift and nonsense variants in the last exon producing C-terminally truncated proteins, argues against loss-of-function as the primary pathogenic mechanism, as additional loss-of-function variants elsewhere in the gene would be expected. Instead, functional analyses revealed two distinct pathogenic processes: partial loss-of-function (LoF) effect for truncating variants and gain-of-function (GoF) effect for missense and in-frame deletions. The absence of early truncating or null alleles (i.e. variants introducing premature stop codons in nonsense-mediated decay sensitive regions) suggest embryonic lethality, reflecting the essential role of RBMX in development. However, pathogenic mechanisms must be interpreted in light of redundancy with its closest retrocopy, *RBMXL1*. Although *RBMXL1* has historically been considered a nonessential or semi-functional retrogene, our data indicate that it is expressed in the developing cortex and interacts with overlapping RNA and protein targets as *RBMX*. The fact that both human and mouse *RBMXL1,* which arose from independent retrotransposition events, converge on a shared neurodevelopmental function suggests evolutionary pressure to preserve RBMX-like activity. This supports the broader hypothesis that retrocopies offer a flexible mechanism for maintaining critical gene dosage.

We further demonstrated that RBMX variants impact the delamination of neuronal cells, reflecting defects in proliferation and/or early neuronal differentiation, although additional cellular phenotypes such as alterations in neuronal positioning or connectivity, cannot be excluded. In mice, delamination assays highlighted a domain-specific dichotomy in RBMX-related pathological mechanisms: variants within the conserved N-terminal RRM domain exert dominant effects, whereas C-terminal truncating variants lead to partial LoF, likely by disrupting splicing regulation, including *ATRX* exitron formation, and potentially through altered RBMXL1 interaction. This partial LoF can be buffered by RBMXL1, emphasizing the underappreciated concept that compensatory expression of the retrocopy can modulate the phenotypic expressivity of RBMX variants. Enhancing RBMXL1 expression or activity may thus represent a rational therapeutic strategy.

The partial functional compensation provided by *RBMXL1* mirrors the well-known interplay between *SMN1* and *SMN2* in spinal muscular atrophy (SMA). In SMA, the presence of *SMN2*, an almost identical paralog of *SMN1*, can partially rescue motor neuron function, and the number of *SMN2* copies correlates inversely with disease severity.^24,25^ *SMN1* and *SMN2* differ mainly by a single nucleotide in exon 7, causing *SMN2* to predominantly produce a truncated, less functional protein due to exon skipping. This splicing difference is targeted by therapies like nusinersen, which promote exon 7 inclusion in *SMN2* transcripts to restore full-length, functional SMN protein.^26^ Similarly, *RBMXL1* can substitute for RBMX essential function in a *RBMX* pathological context. However, like *SMN2*, *RBMXL1* is not fully functionally equivalent to its ancestral gene, leading to incomplete rescue. In addition, its compensation may be stage- or cell type- restricted, as *RBMXL1* expression varies in progenitors versus neurons.

Beyond *RBMX* and *RBMXL1*, several other hnRNP family members also have retrocopies that may contribute to functional redundancy. Examples include HNRNPA1L2^27^ and HNRNPH2, HNRNPF, ^28^ HNRNPH1P3^27^ originated from ancient retrotransposition events involving HNRNPA1 and HNRNPH1, respectively. These examples reflect a broader pattern in which several hnRNP genes possess processed pseudogenes or retrocopies with variable expression, which functional impact remains underexplored. Our findings illustrate how RBMX/RBMXL1 compensation modulates phenotype and point to the broader need for systematic investigation of retrogene expression and function as potential disease modifiers

## METHODS

### Inclusion and ethics statement

This study was conducted in accordance with the ethical standards of all participating countries. Informed consent was obtained from the parents or legal guardians of all patients included in the study. Upon receipt at University Hospital Essen or the respective participating center, patient data and samples were pseudonymized for genetic analysis. Information on the patients’ sex (but not gender) was extracted from clinical records. The study was approved by the Ethics Committee of University Hospital Essen (reference 23-11178-BO).

### Patients and *RBMX* variant identification

We initially sequenced the exome of the two brothers and parents of Family 7 at the sequencing facility of ICM (iGenSeq). Exons were captured from fragmented genomic DNA samples using the SeqCap EZ Solution-Based Enrichment v3 (Roche), and paired-end 75-base massive parallel sequencing was carried out on an Illumina NextSeq500. Bioinformatics analyses were performed using the in-house pipeline, as previously reported.^29^ Patients from families 3 and 6 were subsequently diagnosed through an NGS panel targeting genes implicated in agenesis of the corpus callosum (“callosome”), in which *RBMX* had been included as a candidate gene.^30^ The p.Arg381* variant in Family 9 was identified by whole-exome sequencing (WES) performed at Hôpital Necker. Patient 5 underwent research-based WES, and the p.Gln345* variant was discovered through a collaboration within the IRC^5^ consortium (https://www.irc5.org/). Patient 8 was identified through the DECIPHER database (DECIPHER Patient 488393)^31^ and patients 1, 2, 4 were identified through GeneMatcher^32^ as part of data sharing and matchmaking efforts. Variants identified by next-generation sequencing (exome or panel) were validated and analyzed in parents or relatives by Sanger sequencing. The predicted effect of mutations was interpreted with Alamut 2.11 (Interactive Biosoftware).

### Phylogenetic analyses of *RBMX* and *RBMX*-like genes

Orthologues of *RBMX*, *RBMXL1* and *RBMXL2* (from human) and *Rbmxl1* and *Gm7324* (from *Mus musculus*) were obtained from Ensembl using the Comparative Genomic tools^33^. The coding sequences of the queried genes, their orthologues and *Rbmx* from zebrafish (to be used as outgroup), as well as the number of exons from known transcripts, were retrieved from available genomes using biomaRt^34,35^. In case of multiple transcripts, only one was kept based on the following criteria: 1) the transcript with “MANE Select” and “GENCODE basic” flags for *Homo sapiens*, 2) the transcript with “GENCODE basic” flag for *Mus musculus*, 3) the transcript with the longest coding sequence. Sequences that fulfilled the following criteria were kept for further analyses: a) no sequence ambiguities, b) corresponding genome with chromosome assembly-level as stated by ENA (genomes from hybrid species were excluded; when more than one genome per species was available, the one with higher scaffold N50 was kept) and c) translated protein sequence containing the amino acids RDGYG motif, which is conserved up to zebrafish and located 11 amino acids upstream the region necessary for RNA-binding. Assignment of sequences to one of the six gene groups was, when possible, based on the indication of confidence of orthology from Ensembl: *RBMX*, high confidence orthologue (hco) of human *RBMX* only or, hco of human *RBMX* and *RBMXL1* and more than six exons; *RBMXL1*, hco of human *RBMXL1* only; *RBMXL2*, hco of human *RBMXL2* only; *RBMXL1a*, hco of mouse *Rbmxl1a* only; *RBMXL1b*, hco of mouse *Rbmxl1b* only; retrocopy, three exons or less. Genes for which none of these conditions was fulfilled or genes named as retrocopies were manually curated, considering the percentage of identity with each of the five genes used to query Ensembl and the number of exons (retrocopy: one coding exon; RBMXLg: multiple coding exons). Multiple alignment of the remaining 83 DNA coding sequences was performed using ClustalW and default parameters in Geneious Prime 2019.2.3 software (Biomatters Ltd.). Phylogenetic tree was generated using Geneious TreeBuilder with Tamura-Nei genetic distance model, neighbor-joining tree-building method and zebrafish *Rbmx* as outgroup.

### Analysis of *RBMX* and *RBMXL1* specific expression

#### In human brains using Brainspan

To evaluate the specific expression of *RBMX* and *RBMXL1*, we aligned and compared mRNA isoforms NM_002139.3 (*RBMX*) and NM_001162536.2 (*RBMXL1*) using CLUSTAL Omega v1.2.4. This analysis revealed three possible specific loci (*RBMX*: chrX:136880722– 136880780; *RBMXL1*_1: chr1:88979457–88981948 (shown in **Fig. 4b**); *RBMXL1*_2: chr1:88983972–88984066). We assessed short-read mappability by aligning simulated 50-mer and 100-mer sequences to the human genome. The high uniqueness at both lengths supports confident mapping at a read length of 75 bp. Expression levels for each locus were quantified using both RPKM and raw read counts in data from Brainspan (http://brainspan.org), and results were visualized through locus-specific coverage plots.^17^

#### In human brains and cortices using CZ CELLxGENE Discover platform

To explore the expression pattern of *RBMX* and *RBMXL1* across brain regions, developmental stages and cell types, CZ CELLxGENE (https://cellxgene.cziscience.com/) datasets were used.^18^ Metadata and expression matrices were filtered to include only primary, normal brain samples with annotated developmental stages and cell types. Standardized names for brain regions and cell types were generated using custom mapping tables, and only cell types and developmental stages passing manual curation were retained for analysis. From initial census data, samples were filtered through the following criteria: (1) primary data only (is_primary_data=True), (2) normal tissue (disease=’normal’), (3) brain tissue (tissue_general=’brain’), (4) known developmental stage (development_stage≠’unknown’), and (5) manually curated cell types and brain regions passing quality assessment. This filtering approach ensured data quality while maintaining biological relevance across developmental stages.Expression data for *RBMX* (ENSG00000147274) and *RBMXL1* (ENSG00000213516) were extracted from the census using the cellxgene_census Python API. For each gene, only datasets in which the gene was detected were included (*RBMX*: 149 datasets for brain regions, 97 for cortex; *RBMXL1*: 115 datasets for brain regions, 66 for cortex). Cells were grouped by standardized brain region or cortex cell type (x-axis) and developmental stage (y-axis). Gene expression was considered positive when normalized expression values exceeded 0. Expression calculations were performed only among expressing cells to avoid bias from zero-inflation inherent in single-cell RNA-seq data.

For each gene, within each group (region/cell type × developmental stage), the percentage of cells expressing the gene (expression > 0) and the cell-weighted average normalized expression among expressing cells were calculated. The cell-weighted average was computed by: (1) processing each dataset separately to calculate expression metrics per cell group, (2) summing expression values and cell counts across all contributing datasets, and (3) dividing the total expression sum by the total number of expressing cells across all datasets. To enable direct visual comparison between genes, expression values were jointly scaled: the maximum cell-weighted average expression observed for either gene across all groups was used as the reference, and all group means were divided by this value. Expression values were log-normalized within the CellxGENE Census pipeline to account for technical variation between datasets. This normalization approach mitigates batch effects while preserving biological signal across the diverse dataset collection. All analyses were performed on a High-Performance Computing Cluster using Python 3.11.12 with the following key packages: cellxgene_census 1.17.0 (API access), scanpy 1.11.1 (single-cell analysis), pandas 2.2.2 (data manipulation), matplotlib 3.10.3 (visualization), and numpy 2.2.6 (numerical operations).

#### In progenitors and neurons from mouse and human single-cell transcriptomic data

The expression matrices and the cell group annotation matrices, which indicates the classification of individual cells based on transcriptomic similarities, were retrieved from published datasets.^19,23^ Expression values for the *RBMX*/*Rbmx* and *RBMXL1/Rbmxl1* genes were extracted from these matrices. By convention, each row of the expression matrix corresponds to a gene and each column to a single cell, with each entry representing the expression level of a given gene in a particular cell. Based on these data, the number of progenitor or neuronal cells expressing both genes, only one of the two genes, or neither gene was quantified.

### Immunolabeling on slices of fixed-tissues for human fetuses

Human fetal brain experiments were approved by the Comité d’Ethique Hospitalo-Facultaire Universitaire de Liège (EudraCT: 2017-002366-45). The human fetal brain sample was fixed with 4% PFA overnight at 4°C, followed by dehydration with 30% sucrose. The samples were then cryosectioned at a thickness of 20 µm. The primary RBMX antibody (Cell signaling #14794) was diluted at a 1:200 ratio and incubated overnight at 4°C. Secondary antibody was diluted at a 1:500 ratio and incubated 4 hours at 4°C (ThermoFisher Scientific #A-11055 and #A-31572) and nuclear counterstain (DAPI, Sigma #D9542). Images were taken with Zeiss LSM980 20x objective. The sex information for the human fetal brain sample was not available.

### Cell culture

Mouse neuroblastoma N2A (ATCC) cells were cultured in DMEM (Gibco) supplemented with 5% Fetal calf serum (FCS) and gentamicin 40 μg/mL in a humidified atmosphere containing 5% CO2 at 37°C. Human embryonic kidney 293 and 293T (HEK293 (DSMZ), HEK293T (ATCC)) cells were cultured in Dulbecco’s modified Eagle’s medium (DMEM, Gibco) with 10% FCS, 100 U/mL penicillin and 100 μg/mL streptomycin (Gibco). Human neural stem cells (hNSCs) with the genetic background GM01869 (male) were obtained from I-Stem and have been previously described.^36,37^ hNSCs were seeded on poly-ornithine-and laminin-coated dishes and maintained in N2B27 medium consisting of a 1:1 mixture of DMEM/F12 and Neurobasal medium (ThermoFisher Scientific), supplemented with N2 and B27 supplements (ThermoFisher Scientific), 2-mercaptoethanol (ThermoFisher Scientific), brain-derived neurotrophic factor (BDNF, 20 ng/mL), fibroblast growth factor-2 (FGF-2, 10 ng/mL; both from PeproTech), epidermal growth factor (EGF, 10 ng/mL; R&D Systems), and gentamicin (100 µg/mL; Sigma). The culture medium was replaced every two days. For neuronal differentiation, hNSCs were seeded at a density of 15,000 cells/cm² and initially maintained for 2 days in complete medium containing BDNF, EGF, FGF-2, and gentamicin. After two days, the medium was replaced with differentiation medium consisting of the same base medium supplemented only with BDNF and gentamicin. Cells were maintained under these differentiation conditions for 19 days, with medium changes every two days.

Patient fibroblasts cultivated from skin biopsies were available for three patients. The following age-matched fibroblast lines were obtained from the Gerontology Research Center (GRC) Cell Culture Collection | Baltimore Longitudinal Study on Aging (BLSA) and the NIGMS Human Genetic Cell Repository the at the Coriell Institute for Medical Research: AG06234, GM03348, GM08398, GM02037. All fibroblasts cell lines were cultured in DMEM (Gibco) supplemented with 10% FCS (Gibco) and 100 U/mL penicillin and 100 μg/mL streptomycin (Gibco) and maintained at 37°C in a humidified atmosphere with 5% CO₂. Culture medium was replaced every 2–3 days.

### Cloning and plasmid constructs

Wild-type (WT) human *RBMX* coding sequence (CDS) corresponding to RefSeq: NM_002139.3 was obtained from Sino Biological (Ref HG16560-UT) and subcloned by restriction-ligation into the pCAG (Addgene, Plasmid #11160) vector. Constructs carrying the human *RBMX* variants, with or without a HA tag at the N-terminal position, including c.37G>C (p.G13R), c.166T>C (p.F56L), c.247_249del (p.Q83del), c.484_486del (p.P162del), c.1033C>T (p.Q345*), c.1037_1059del (p.E346Gfs*9), c.1057_1058del (p.M353Gfs*9), c.1063dup (p.R355Kfs*8), and c.141C>T (p.R381*) were created from the WT CDS by Sequence and Ligation Independent Cloning (SLIC). WT human *RBMXL1* coding sequence (RefSeq: NM_001162536.3) was amplified by nested PCR from genomic DNA of a healthy control individual and subcloned by restriction-ligation into the pCAG (Addgene, Plasmid #11160). Constructs carrying the WT human *RBMXL1* CDS, with a HA or FLAG tags at the N-terminal position were created from the WT CDS by SLIC. The lentiviral construct pLV-CAG empty vector was generated by subcloning NdeI/BsrGI fragments from pCAG empty vector (Addgene, Plasmid #11160) as donor plasmids into the pLV-CMV-mCherry-WPRE vector (Addgene, Plasmid #36084). Final pLV constructs expressing FLAG-tagged WT RBMX or WT RBMXL1 were generated by transferring XbaI/BsrGI fragments from the previously described pCAG-FLAG-RBMX and pCAG-FLAG-RBMXL1 plasmids into the pLV-CAG empty vector backbone. All constructs were fully sequenced to confirm the absence of mutations introduced during the cloning process.

As the high degree of homology between *RBMX* and *RBMXL1*, which made it difficult to use standard tools to identify unique target sequences, miRNAs targeting the CDSs of mouse *Rbmx* (NM_011252.5) and mouse *Rbmxl1* (NM_001252089.1) were manually designed by aligning both sequences (**Supplementary Table 11**). Sense and antisense oligos (**Supplementary Table 12**) were annealed and the resulting duplex was subcloned in pCAG-mir30 (Addgene, Plasmid #14758)^38^ vector digested with XhoI and EcoRI. pCAG:H2B-eGFP was obtained from Addgene (Plasmid #32599). Plasmid DNAs for IUE experiments were purified using the Endotoxin-Free Plasmid Purification Kit (Macherey-Nagel), while plasmids used in all other experiments were prepared with the same kit but without the Endotoxin-Free quality.

### RNAseq analysis

For RNA-seq experiment from N2A cells, transfections were performed at 50-60% confluency using Lipofectamine 2000 (ThermoFisher Scientific), according to the manufacturer’s protocol. N2A cells were co-transfected in one well of a 6-well plate per condition with 1 μg of pCAGGs-linkB-IRES-GFP and either 3 μg of pCAG-miR30-scramble or a pool of pCAG-miR30-miRNAs targeting mouse *Rbmx* (*Rbmx* #1 + #2 + #3) or *Rbmxl1* (*Rbmxl1* #1 + #2) (1μg each). A total of 3 μg of miRNA constructs was used per condition. 48 hours post-transfection, N2A cells were washed with 1 mL of PBS 1X, treated with 1 mL of diluted trypsin for 1-2 min, and dissociated by pipetting. Trypsin was neutralized by adding 1 mL of DMEM (1 g/L glucose) supplemented with 5% FCS and 40 μg/mL gentamicin. N2A cells were centrifuged (5 min, 1200 rpm), and resuspended in cold FACS buffer (PBS 1X, 0.5% BSA, 2 mM EDTA). N2A cells were then passed through a 100 μm cell strainer and kept on ice until sorting of GFP-positives cells. N2A cells were sorted using FACS ARIA II (BD) with a 100 µm nozzle, in cold FACS buffer. After sorting, N2A cells were centrifuged (5 min, 1000 rpm, 4 °C), the supernatant was removed, and cell pellets were snap-frozen in liquid nitrogen before storage at −80 °C until RNA extraction. Total RNA were extracted using the AllPrep DNA/RNA Kits from Qiagen. mRNA libraries were prepared using the ILLUMINA Stranded Total RNA Prep, Ligation with Ribo-Zero Plus reagents, following manufacturer’s recommendations. Final pooled library samples were sequenced on ILLUMINA NovaSeq6000 with S1-200 cartridge (2x1600Millions 100 bases reads), corresponding to 2x100Millions reads per sample after demultiplexing. For RNA-seq analysis from N2A cells experiment, reads were preprocessed to remove adapter, polyA and low-quality sequences (Phred quality score below 20) using cutadapt v.1.10^39^ and then, reads shorter than 40 bases were discarded for further analysis. Reads were mapped onto the mm10 assembly of *Mus musculus* genome using STAR v.2.5.3a.^40^ Quantification of gene expression was performed from uniquely aligned reads using HTSeq-count v.0.6.1p1^41^, with union mode and annotations coming from Ensembl version 102. Comparisons of interest have been performed using R v.3.3.2 with DESeq2 v.1.16.1^42^. More precisely, read counts were normalized from the estimated size factors using the median-of-ratios method and a Wald test was used to estimate the *P* values. *P*-values were adjusted for multiple testing using the Benjamini and Hochberg method.^43^

For RNA-seq experiments from patients’ fibroblasts, cells were cultured in AmnioMAX (Thermo Fisher Scientific). Total RNA was extracted using the AllPrep DNA/RNA/miRNA Kit (Qiagen, ref. 80224) and mRNA libraries were prepared with the Illumina Stranded mRNA Kit and sequenced on an Illumina NextSeq 500 using a High Output 300-cycle kit. FASTQ files were generated with Illumina’s bcl2fastq software, and reads were aligned to the hg38 reference genome using the snakemake pipeline (STAR aligner) available at the link https://zenodo.org/records/7885540.^44^

### iCLIP Data Processing and Peak Calling

High-confidence RNA-protein crosslink sites from RBMX-FLAG expressing Flp-In HEK293 cells were obtained from GSE233498.^3^ For this reanalysis, single-nucleotide resolution crosslink sites from three biological replicates were further processed using iCount v2.0 with human genome reference GRCh38 and Ensembl annotation release 88. Peak calling was performed independently for each replicate using a 10-nucleotide half-window and grouped by Ensembl gene_id. Peaks were identified at a false discovery rate (FDR) of 0.05 using 1,000 permutations. Only peaks reproducible in at least two of three replicates were retained for downstream analysis. Reproducible peaks were annotated using iCount with Ensembl gene models to assign genomic features including coding sequences (CDS), untranslated regions (UTRs), introns, and non-coding RNAs (ncRNA). For peaks present in multiple replicates, the highest score was retained as the representative binding strength. RNA-seq data from the same cell line (GSM7429573) were processed using the nf-core/rnaseq pipeline v3.19.0 with STAR-RSEM quantification. Genes with transcript-per-million (TPM) values > 1 were considered expressed. Only peaks mapping to expressed genes were included in the final analysis to focus on transcriptionally active loci. For each gene, peak counts and cumulative scores were calculated across genomic features. Genes were ranked by total peak number, and Ensembl gene IDs were mapped to official gene symbols using GTF annotation for biological interpretation.

### *SMN2* splicing assay

Alternative splicing of *SMN2* exon 7 was assessed using a pCl-SMN2 splicing minigene containing human *SMN2* exons 6 to 8. pCI-SMN2 was a gift from Elliot Androphy (Addgene plasmid # 72287 ; http://n2t.net/addgene:72287 ; RRID:Addgene_72287).^45^ HEK293 (DSMZ) were seeded at a density of 62,500 cells/cm² per well of a P6-well plate and transfected using jetOPTIMUS® DNA Transfection Reagent (Polyplus) with a 1:1 ratio of DNA/polymer according to the manufacturer’s protocol. Cells were transfected with 1 µg of pCl-SMN2 and 0.5 µg of pCAGEN empty vector (negative control) or a pCAGEN vector containing the CDS for human wildtype or mutant *RBMX* or *RBMXL1* with an N-terminal HA-tag. 24 hours post-transfection, total RNA was extracted using the Maxwell® RSC simplyRNA Cells Kit (Promega) and the Maxwell® RSC Instrument (Promega) according to the manufacturer’s protocol. 1 µg of extracted RNA was submitted to DNase treatment (RQ1, Promega) and used for synthesis of cDNA using the LunaScript® RT SuperMix Kit (NEB) following the manufacturer’s instructions. To amplify both FL-*SMN2* and *SMN2Δex7*, 2 µl of cDNA were used for PCR using the HotStarTaq Master Mix Kit (1000U, Qiagen), the pCl backbone-specific forward primer pCl-fwd and reverse primer SMNex8-rev from Moursy et al.^21^ Amplification of *HPRT* was done using primers from Vezain et al.^46^ was used to check the cDNA quality. Both PCRs were run at 58°C with 22 cycles (*SMN2*) or 25 cycles (*HPRT*) and PCR products were separated by agarose gel electrophoresis for 45 min at 150V. Gels were imaged using a ChemoStar Touch (Intas) with UV transilluminator. For quantification of FL-*SMN2* to *SMN2Δex7* ratio, cDNA was diluted 1:2 with ddH_2_O and amplified by RT-qPCR using the KAPA SYBR® FAST qPCR Master Mix (2X) Kit (KAPA BIOSYSTEMS) in a LightCycler 480 (Roche). The pCl backbone-specific forward primer pCl-fwd was combined with reverse primer SMN2-ex6_7_8R to amplify FL-*SMN2* and with SMN2-ex6_8R to amplify *SMN2Δex7*. The qPCR was run with the supplier’s standard program with 60°C annealing temperature. All primer sequences are available in **Supplementary Table 13**.

### *ATRX* splicing assay

Alternative splicing of endogenous *ATRX* exon 9 was assessed in HEK293 (DSMZ) cells. Cells were seeded at a density of 52,600 cells/cm² per well of a P24-well plate. Knockdown of endogenous *RBMX* expression was induced by transfecting 25 pmol of a custom siRNA targeting *RBMX* 3’UTR (Dharmacon, Sense: 5’-AAGUAAAAGUAUCCCCUAAUU-3’, Antisense: 5’-UUAGGGGAUACUUUUACUUUU-3’) or non-targeting control siRNA (Dharmacon, D-001210-01-05) using Lipofectamine RNAiMAX Transfection Reagent (ThermoFisher Scientific) according to the manufacturer’s protocol. 24 hours after knockdown, the *ATRX* splicing phenotype was rescued by transfection with 0.5 µg of pCAGEN empty vector (negative control) or a pCAGEN vector containing the CDS for human wildtype *RBMX* or *RBMXL1* with an N-terminal HA-tag. Transfection was carried out using jetOPTIMUS® DNA Transfection Reagent (Polyplus) with a 1:1 ratio of DNA/polymer according to the manufacturer’s protocol. 24 hours post rescue, total RNA was extracted using the Maxwell® RSC simplyRNA Cells Kit (Promega) and the Maxwell® RSC Instrument (Promega) according to the manufacturer’s protocol. 1 µg of extracted RNA was submitted to DNase treatment (RQ1, Promega) and subsequently utilized for synthesis of cDNA using the LunaScript® RT SuperMix Kit (NEB) in accordance to the manufacturer’s instructions. To amplify both *ATRX* exon 9 full length and *ATRX* with exitron, 2 µl of cDNA were used for PCR using the HotStarTaq Master Mix Kit (1000U, Qiagen) with forward primer ATRX_exon9-fwd1 and reverse primer ATRX_exon9-rev at 60°C for 35 cycles. Amplification of *HPRT* to verify cDNA quality, PCR product separation and imaging was carried out as described for *in-vitro* splicing analysis of *SMN2* with 35 cycles. PCR products were separated by agarose gel electrophoresis for 45 min at 150V. Gels were imaged using a ChemoStar Touch (Intas) with UV transilluminator. To quantify the ratio of FL-*ATRX* to all *ATRX* isoforms containing exon 9, cDNA was diluted 1:20 with ddH_2_O and amplified by RT-qPCR using the LightCycler 480 SYBR Green I Master in a LightCycler 480 (both from Roche). Primers ATRX_exon9-fwd1 and ATRX_exon9-rev2 were combined to amplify *ATRX* full-length exon 9 and primers ATRX_exon9-fwd2 and ATRX_exon9-rev3 were combined to amplify all *ATRX* isoforms containing exon 9. The qPCR was run with the supplier’s standard program with 60°C annealing temperature, an annealing duration of 45 sec and an elongation duration of 25 sec. All primer sequences are available in **Supplementary Table 13**.

### Quantification of *ATRX* full-length and short isoform by RT-qPCR

For hNSCs, cells were seeded at a density of 17,000 cells/cm² in a one well of P24-well plate and transfected using Lipofectamine LTX (ThermoFisher Scientific) according to the manufacturer’s protocol. hNSCs were transfected with 25 pmol of either control siRNAs (Dharmacon, D-001810-10-05) or *RBMX/RBMXL1*-targetting siRNAs (Dharmacon, L-011691-01-0005). 48 hours post-transfection, total RNA was extracted using TRIzol reagent (Thermo Fischer Scientific) from transfected cells, following the manufacturer’s protocol. All RNA samples were submitted to DNAse treatment (Turbo DNAse, ThermoFisher Scientific). cDNA samples were synthetized with SuperScript IV Reverse Transcriptase (ThermoFisher Scientific, 18090200) and PCR was performed using the following *ATRX* primers with 16 touchdown cycles (70°C to 54°C) followed by 21 cycles at 59°C. PCR products were analyzed using a 2100 Bioanalyzer (Agilent Technologies). The same protocol for RNA extraction, DNase treatment, cDNA synthesis, and PCR was applied to non-transfected samples (fibroblast lines and hiPSCs).

For fibroblast lines, cells were seeded at a density of 41,700 cells/cm² in two wells of 24-well plate and transfected using Lipofectamine 3000 (ThermoFisher Scientific), following the manufacturer’s protocol. Fibroblasts were transfected with 2.5 µg of pLV-CAG empty vector or a pLV-CAG vector containing the coding sequence for human wildtype *RBMX* or *RBMXL1* with an N-terminal FLAG-tag and co-expressing mCherry. At 36 hours post-transfection, fibroblasts were washed with 1 mL of 1X DPBS (Gibco), treated with 1 mL of diluted trypsin for 1-2 min, and dissociated by pipetting. Trypsin was neutralized by adding 1 mL of DMEM (1 g/L glucose) supplemented with 10% FCS (Gibco), and 100 U/mL penicillin, and 100 μg/mL streptomycin (Gibco). Fibroblasts were centrifuged (5 min, 600 rpm), and resuspended in cold FACS buffer (PBS 1X, 0.5% BSA, 2 mM EDTA). Fibroblasts were then passed through a 100 μm cell strainer and kept on ice until sorting of mCherry-positives cells. Fibroblasts were sorted using FACSAria Fusion (BD) with a 100 µm nozzle, in cold FACS buffer. After sorting, fibroblasts were centrifuged (5 min, 1000 rpm, 4 °C), the supernatant was removed, and cell pellets were snap-frozen in liquid nitrogen before storage at −80 °C until RNA extraction. Total RNA was extracted using TRIzol reagent (Thermo Fischer Scientific) from transfected fibroblasts, following the manufacturer’s protocol. All RNA samples were submitted to DNAse treatment (Turbo DNAse, ThermoFisher Scientific). cDNA samples were synthetized with SuperScript IV Reverse Transcriptase (ThermoFisher Scientific, 18090200) and PCR was performed using *ATRX* primers with 16 touchdown cycles (70°C to 54°C) followed by 21 cycles at 59°C. PCR products were analysed using a 2100 Bioanalyzer (Agilent Technologies). The linear amplification within the exponential phase of the *ATRX* amplicon on the 2100 Bioanalyzer (Agilent Technologies) was confirmed allowing quantification. All primer sequences are available in **Supplementary Table 13.**

### Yeast two hybrid screen

Yeast two-hybrid (Y2H) interaction screens were conducted by Hybrigenics Services (Paris, France) using full-length human wild-type RBMX (RBMX WT), human full-length RBMXL1, and a truncated human RBMX construct encompassing amino acids 1-380, mimicking the pathogenic p.Arg381* variant. The initial screen was performed using an N-terminal LexA-RBMX-C fusion construct; however, as the LexA-based screen yielded only 20 positive clones, the bait was subsequently transferred to a Gal4 vector to initiate a second screen. In the end, all three bait constructs were cloned in-frame with the Gal4 DNA-binding domain into a yeast expression vector and verified by sequencing. The Gal4-RBMX fusions were used in high-throughput Y2H screens against the Fetal Brain_RP2 library pre-transformed into yeast strains expressing the Gal4activation domain. None of the baits were toxic or auto-activating and the screen was performed the screen without 3-Aminotriazol. Approximately 100 million interactions were tested and analysed. Interacting prey proteins were selected based on growth on selective media and activation of reporter genes. Positive clones were sequenced to identify interactors, and interactions were assigned confidence scores by Hybrigenics’ proprietary Predicted Biological Score (PBS) algorithm. Comparative analysis across the three baits enabled assessment of interaction profiles specific to RBMX WT, RBMXL1, and the RBMX 1–380 truncated form.

### Generation and primary characterization of the *Rbmx^tm2a(KOMP)Wtsi^* knockout mouse

The *Rbmx^tm2a(KOMP)Wtsi^* mouse was generated at the Wellcome Sanger Institute on a pure C57BL/6NTac background by homologous recombination in embryonic stem cells using the knockout-first allele method.^47^ In this strategy, exon 4, common to all transcripts, was targeted, and a LacZ cassette inserted upstream, producing the *Rbmx^tm2a(KOMP)Wtsi^*knockout-first allele (reporter-tagged insertion with conditional potential) (**Fig. 3a**). The LacZ cassette provides the advantage of enabling gene expression studies.

Through collaboration with the Sanger Institute Mouse Genetics Project, we received brain samples from *Rbmx^tm2a(KOMP)Wtsi^* mice and performed comprehensive neuroanatomical studies on three hemizygous mutants and 498 baseline WT mice at 16 weeks of age, as previously described.^16,48^ A validated statistical model (G*Power) was applied to detect neuroanatomical defects with an effect size ≥10% at 80% power.^15^ Brains were fixed in 4% buffered formalin for 48 hours, transferred to 70% ethanol, and paraffin-embedded. Coronal sections (5-μm thickness) were obtained at Bregma +0.98 mm, –1.34 mm and –5.80 mm (Allen Mouse Brain Atlas) using a Leica RM 2145 microtome. Sections were double-stained with 0.1% Luxol Fast Blue (Solvent Blue 38; Sigma-Aldrich) and 0.1% Cresyl violet acetate (Sigma-Aldrich), then scanned at 20× resolution using the Nanozoomer whole-slide scanner 2.0HT C9600 series (Hamamatsu Photonics, Japan). Images were quality controlled for sectioning accuracy, asymmetries, and histological artefacts. All samples were assessed for cellular ectopia (misplaced neurons). Seventy-eight brain parameters (area and length) were measured blind to the genotype across the three coronal sections as described in ^48^ (**Supplementary Tables 3-4**). Data were analyzed using a t-test to determine whether a brain region is associated with neuroanatomical defect or not.

Quality control of head orientation during X-ray (cephalometrics) was established using three landmarks on each side of the head. Seven mutants were compared to 14 local controls and 11 distances were taken (**Supplementary Table 5**) which are summarized in **Supplementary Fig. 4e** and were taken blind to the genotype. Data were analyzed using a t-test of equal variance to determine whether a measurement is significantly associated with a defect or not.

For LacZ staining (**Supplementary Fig. 4c**), tissues were fixed in 2% paraformaldehyde/0.2% glutaraldehyde in PBS with 2 mM MgCl₂. Whole-mount tissues were incubated in X-gal staining solution (1 mg/mL X-gal, 5 mM potassium ferri/ferrocyanide, 2 mM MgCl₂, 0.02% NP-40, 0.01% sodium deoxycholate in PBS, pH 7.4) at 37 °C until blue precipitate was visible (2–16 h). Samples were post-fixed in 4% PFA and imaged. Staining was scored as positive (+), absent (–), or non-specific (NS).

### Mice

All animal studies were conducted in accordance with French regulations (EU Directive 86/609 – French Act Rural Code R 214-87 to 126) and all procedures were approved by the local ethics committee and the Research Ministry (APAFIS#15691-201806271458609 and #44708-2023090108462873). Mice were bred at the IGBMC animal facility under controlled light/dark cycles, stable temperature (19°C) and humidity (50%) condition and were provided with food and water ad libitum. Timed-pregnant WT CD1 (Charles River Laboratories) females were used for RT-qPCR, Western blot, immunofluorescence analyses, and *in utero* electroporation experiments with the different constructs at embryonic day 13.5. All the experiments have been conducted in males. The sex was identified as follow: *Sry* genotyping using the forward primer (5’-GTCACAGAGGAGTGGCATTT-3’) and reverse primer (5’-AGTCTTGCCTGTATGTGATGG-3’), or by gonad dissection.

### *In utero* electroporation

In utero electroporation (IUE) was performed as described previously^49,50^. Briefly, CD1 pregnant females were anesthetized with isoflurane (2L/min of oxygen; 4% isoflurane in the induction phase followed by 2% isoflurane during surgery; Tem Sega). The uterine horns were exposed, and a lateral ventricle of each embryo was injected using pulled glass capillaries (Harvard apparatus, 1.0OD*0.58ID*100mmL) with Fast Green (1 μg/μl; Sigma) combined with different amounts of DNA constructs using a micro injector (Eppendorf Femto Jet). Plasmids were electroporated into the neuronal progenitors adjacent to the ventricle by 5 electric pulses (35V) for 50 ms at 950 ms intervals using electrodes (diameter 3 mm; Sonidel CUY650P3) and ECM-830 BTX square wave electroporator (VWR international). After electroporation, embryos were placed back in the abdominal cavity and the abdomen was sutured using surgical needle and thread. For E15.5 analysis, pregnant mice were sacrificed by cervical dislocation 2 days after surgery. Only males were further analysed. Conditions of IUE with plasmids and concentration used are summarized in **Supplementary Table 14.**

### Mouse brain fixation, cutting and immunohistochemistry

E13.5 and E15.5 male embryos were sacrificed by head sectioning and brains were fixed in 4% paraformaldehyde (PFA, Electron Microscopy Sciences) diluted in phosphate buffered saline (PBS, HyClone) overnight at 4°C. For *Rbmx/Rbmxl1* expression analyses, coronal and sagittal 14 μm (E13.5) and 16 μm (E15.5) cryosections were used. For IUE analyses, 16 μm cryosections were prepared from E15.5 brains. All cryosections were subjected to immunolabeling as follows: after fixation, brains were rinsed and equilibrated in 20% sucrose in PBS overnight at 4°C, embedded in Tissue-Tek O.C.T. (Sakura), frozen on dry ice, cut coronally at the cryostat (16 μm thickness, Leica CM3050S) and maintained at −80°C until immunolabeling. For immunolabeling cryosections were permeabilized and blocked with blocking solution (5% Normal Donkey Serum (NDS, Dominic Dutscher), 0.5% Triton-X-100 in PBS) for one hour at room temperature (RT). Sections were then incubated with primary antibodies (**Supplementary Table 15**) diluted in blocking solution overnight at 4°C and with secondary antibodies (**Supplementary Table 15**) and DAPI (dilution 1/1000, 1mg/mL Sigma) diluted in PBS with 0.1% Triton X-100 (Sigma) for one hour at RT. Slides were mounted using Aquapolymount mounting medium (Polysciences Inc) for imaging.

### Immunofluorescence

hNSCs were seeded at a density of 17,000 cells/cm² in a one well of P24-well plate on poly-L-ornithine/laminin-coated coverslips and co-transfected with 400 ng of HA-tagged WT RBMX, WT RBMXL1 or various variants along with 100 ng of pCAG-GFP (Addgene, Plasmid #11150). Transfections were performed using Lipofectamine LTX (ThermoFisher Scientific) according to the manufacturer’s protocol. After 24 hours, cells were washed twice with PBS (HyClone) and fixed for 20 minutes at room temperature in 4% paraformaldehyde (PFA, Electron Microscopy Sciences) and 4% sucrose in PBS. After fixation, cells were rinsed twice in PBS, permeabilized for 10 minutes in 0.2% Triton X-100 (Sigma), and blocked for 1 hour at room temperature in PBS containing 3% BSA (Sigma) and 0.02% Triton X-100. Primary antibodies (**Supplementary Table 15**) were applied overnight at 4°C in PBS with 0.2% BSA and 0.02% Triton X-100. Coverslips were placed cell-side down on antibody drops on Parafilm in sealed Petri dishes. After three times 10-minute washes in PBS with 0.02% Triton X-100 and 0.2% BSA, cells were incubated for 2 hours at room temperature with secondary antibodies (**Supplementary Table 15**) diluted in the same buffer. Coverslips were then washed three times 10- minute washes in PBS with 0.02% Triton X-100 and 0.2% BSA and mounted using Aquapolymount mounting medium (Polysciences Inc) for imaging. Images were acquired using a TCS SP8 X (Leica microsystems) confocal microscope using a 3x OIL HC PL APO CS2 objective. For all experiments, a Z-stack of 1 μm was acquired and the image size was 2048x2048.

Fibroblasts were seeded with a density of 39,500 cells/cm² per well of a P24-well plate on high-precision glass coverslips coated with 0.01% poly-L-lysine (Sigma-Aldrich). After 48 hours, cells were washed with 1x DPBS (Gibco) three times and fixed for 10 min at room temperature with 4% methanol-free formaldehyde (Thermo Scientific). After fixation, cells were washed with 1x TBS (Thermo Scientific) three times and incubated in blocking solution containing 1x TBS, 10% normal goat serum (Abcam) and 0.1% Triton-X (Sigma-Aldrich) for permeabilization and blocking for 1 hour at room temperature. Primary antibodies (**Supplementary Table 15**) were applied overnight at 4°C in blocking solution. After three times 10-minute washes in 1x TBS, cells were incubated for 2 hours at room temperature with secondary antibodies (**Supplementary Table 15**) diluted in blocking solution. Coverslips were washed three times for 10 minutes with 1x TBS and incubated with 1 µg/ml DAPI (Thermo Scientific) in 1x TBS for 10 minutes to stain nuclei. Coverslips were washed again three times for 10 minutes with 1x TBS and mounted using Vectashield Antifade Mounting Medium (Vector Laboratories) for imaging. Imaging was done using a LSM 710, AxioObserver Elyra PS.1 (Zeiss) confocal microscope with an alpha Plan-Apochromat 100x/1.46 Oil DIC M27 Elyra objective. For all experiments, the image size was 1024x1024.

### RNA extraction, cDNA synthesis and RT-qPCR

Total RNA was extracted using TRIzol reagent (Thermo Fischer Scientific) from mouse cortices (E12.5 to P21, CD1 and E15.5 *Rbmx*^tm2a^, all males), non-transfected (fibroblast lines and hiPSCs) and transfected cells (N2A, hNSCs, HEK293T), or using the Maxwell® RSC simplyRNA Cells Kit and Maxwell® RSC Instrument (Promega) (fibroblast lines) according to the manufacturer’s protocol. Male embryos and P3 pups were sacrificed by decapitation, while P21 animals were euthanized by cervical dislocation.

For N2A transfection, cells were transfected at 50-60% confluency using Lipofectamine 2000 (ThermoFisher Scientific), according to the manufacturer’s protocol. 48 hours post-transfection, expression of the transfected genes was assessed by RT-qPCR. To evaluate the efficacy of individual miRNAs (IUE experiments) or pooled miRNAs (N2A cells RNA-seq), N2A cells were co-transfected with 1 μg of pCAGGs-linkB-IRES-GFP and 3 μg of either pCAG-miR30-scramble or pCAG-miR30-miRNA targeting mouse *Rbmx* and *Rbmxl1,* either individually or simultaneously. For individually miRNAs validation, each condition included either *Rbmx* (#1 or #2), *Rbmxl1* (#1 or #2), or *Rbmx+Rbmxl1* (#1 or #2). For pooled miRNAs validation, each condition included a combination of *Rbmx* (#1 + #2 + #3) or *Rbmxl1* (#1 + #2). In all conditions, a total of 3 μg of miRNA constructs was used. For hNSCs transfection, cells were seeded at a density of 17,000 cells/cm² in a one well of P24-well plate and transfected using Lipofectamine LTX (ThermoFisher Scientific) according to the manufacturer’s protocol. To assess *RBMX* and *RBMXL1* knockdown efficiency, hNSCs were transfected with 25 pmol of either control siRNAs (Dharmacon, D-001810-10-05) or *RBMX/RBMXL1*-targetting siRNAs (Dharmacon, L-011691-01-0005). 48 hours post-transfection, gene expression was analyzed by RT-qPCR. All RNA samples were submitted to DNAse treatment (Turbo DNAse, ThermoFisher Scientific). cDNA samples were synthetized with SuperScript IV Reverse Transcriptase (ThermoFisher Scientific, 18090200) and RT-qPCR was done with amplified cDNA and SYBR Green Master Mix (Roche) together with 0.1 μM of forward and reverse primers using a LightCycler 480 (Roche). RT-qPCR was performed using the primers listed in **Supplementary Table 13**.

To assess *RBMX* knockdown efficiency and the thereby induced *ATRX* exitron phenotype in HEK293 cells, cells were transfected with 25 pmol of either a custom siRNA targeting *RBMX* 3’UTR (Dharmacon, Sense: 5’-AAGUAAAAGUAUCCCCUAAUU-3’, Antisense: 5’-UUAGGGGAUACUUUUACUUUU-3’) or non-targeting control siRNA (Dharmacon, D-001210-01-05) using Lipofectamine RNAiMAX Transfection Reagent (ThermoFisher Scientific) according to the manufacturer’s protocol. 24 hours after knockdown, cells were transfected with 0.5 µg of pCAGEN empty vector to mimic the conditions used in the *ATRX* splicing assay. Transfection was carried out using jetOPTIMUS® DNA Transfection Reagent (Polyplus) with a 1:1 ratio of DNA/polymer according to the manufacturer’s protocol. After 24 hours, total RNA was extracted using the Maxwell® RSC simplyRNA Cells Kit (Promega) and the Maxwell® RSC Instrument (Promega) according to the manufacturer’s protocol. 1 µg of extracted RNA was submitted to DNase treatment (RQ1, Promega) and subsequently utilized for synthesis of cDNA using the LunaScript® RT SuperMix Kit (NEB) in accordance to the manufacturer’s instructions. To verify RBMX-specific knockdown, *RBMX*- and *RBMXL1*-specific RT-qPCR was done with amplified cDNA and KAPA SYBR® FAST qPCR Master Mix (2X) Kit (KAPA BIOSYSTEMS) together with 0.2 μM of forward and reverse primers using a Lightcycler 480 (Roche). To assess the *ATRX* exitron phenotype upon *RBMX* knockdown, an RT-PCR amplifying both *ATRX* exon 9 full length and *ATRX* with exitron and quantification of the ratio of FL-*ATRX* to all *ATRX* isoforms containing exon 9 by RT-qPCR were performed as described for the ATRX splicing assay. All primer sequences are available in **Supplementary Table 13**.

To analyze endogenous *RBMX* expression in fibroblasts, total RNA was extracted using the Maxwell® RSC simplyRNA Cells Kit and Maxwell® RSC Instrument (Promega) according to the manufacturer’s protocol.1 µg of extracted RNA was submitted to DNase treatment (RQ1, Promega) and used for synthesis of cDNA using the LunaScript® RT SuperMix Kit (NEB) following the manufacturer’s instructions. RT-qPCR was done with amplified cDNA and KAPA SYBR® FAST qPCR Master Mix (2X) Kit (KAPA BIOSYSTEMS) together with 0.2 μM of forward and reverse primers using a Lightcycler 480 (Roche). RT-qPCR was performed using the primers listed in **Supplementary Table 13**.

### CLIP-qPCR

hNSCs were transfected with 2 μg of HA-tagged WT RBMX, WT RBMXL1, or empty vector (pCAG) constructs. After 24 hours, cells from 3 wells of a 6-well plate per condition were pooled, and rinsed once with cold 1x PBS, and then proceeded for UV crosslinking (254 nm, 400 J/cm^2^, Stratalinker) on ice. hNSCs from 3 wells of 6-well plate were lysed in 400 µL of lysis buffer [50 mM Tris·HCl, pH 7.4, 100 mM KCl, 1 mM MgCl_2_, 0.1 mM CaCl_2_, 0.1% SDS, 1% NP40, 0.5% sodium deoxycholate, 30 U of RNasin (Promega), 1x cOmplete EDTA-free protease inhibitor cocktail and 10 U of DNase (Turbo DNAse, ThermoFisher Scientific)] and incubated for 5 min at 37°C. Lysates were spun down 15 min at 13,000 rpm at 4°C, and supernatants were precleared by incubation with 25 μL of free Dynabeads protein G (ThermoFisher Scientific) and then on 25 μL of Dynabeads protein G coupled to mouse anti-rabbit IgGs (2.5 μg) (Jackson ImmunoResearch, 211-002-171) for 1 hour each. The lysates were then incubated overnight with agitation on 25 μL of anti-HA Magnetic beads (88836, Thermo Scientific). After immunoprecipitation, the supernatants were saved for RNA extraction as “inputs” (HA-RBMX, HA-RBMXL1 and empty vector (pCAG)), and the beads were washed three times with 500 µL of high-salt washing buffer [50 mM Tris·HCl, pH 7.4, 1 M KCl, 1 mM EDTA, 1% NP40, 0.5% sodium deoxycholate, and 0.1% SDS; 4 °C]. RNAs from hNSCs transfected with HA-RBMX, HA-RBMXL1, or empty vector (pCAG) constructs were recovered (“CLIP samples”) by treatment of the beads with 0.4 mg of proteinase K in buffer (100 mM Tris·HCl, pH 7.4, 50 mM NaCl, and 10 mM EDTA) for 20 min at 37 °C.

RNA was extracted from “inputs” hNSCs transfected with HA-tagged WT RBMX, WT RBMXL1, or empty vector (pCAG) constructs, as well as from “CLIP samples” corresponding to the same conditions, using phenol/chloroform followed by chloroform/isoamyl alcohol (20:1) extraction and ethanol precipitation in the presence of alcool isopropylique (v/v). Subsequently, total RNA was treated with DNase (Turbo DNAse, ThermoFisher Scientific) and 500 ng of RNA was used for reverse transcription using SuperScript IV (ThermoFisher Scientific, 18090200). qRT-PCR was done with amplified cDNA and SYBR Green Master Mix (Roche) together with 0.1 μM of forward and reverse primers using a Lightcycler 480 (Roche). qRT-PCR was performed on RNA extracted from inputs and CLIP samples using the primers listed in **Supplementary Table 13**.

### Protein extraction

Proteins from mouse cortices (E12.5 to P21, CD1, all males) and transfected cells (HEK293T and hNSCs) were extracted. Male embryos and P3 pups were sacrificed by decapitation, while P21 animals were euthanized by cervical dislocation. For HEK293T transfection, cells at 50-60% confluency were transfected with 2.5 μg of plasmid (HA-tagged WT RBMX, WT RBMXL1, HA-p.R381* or empty vector (pCAG)) using Lipofectamine 2000 (ThermoFisher Scientific), according to the manufacturer’s protocol. To validate the interactome, lysates from 4 wells of a 6-well plate were pooled for immunoprecipitation 24 h post-transfection. For hNSCs transfection, cells were seeded at a density of 17,000 cells/cm² in a one well of P24-well plate and transfected using Lipofectamine LTX (ThermoFisher Scientific) according to the manufacturer’s protocol. To assess *RBMX* and *RBMXL1* knockdown efficiency, hNSCs were transfected with 25 pmol of either control siRNAs (Dharmacon, D-001810-10-05) or *RBMX/RBMXL1*-targetting siRNAs (Dharmacon, L-011691-01-0005). 48 hours post-transfection, protein levels were analyzed by Western blot. Proteins were extracted as follows: cells or tissue were lysed in RIPA buffer (50 mM Tris pH 8.0, 150 mM NaCl, 5 mM EDTA pH 8.0, 1% Triton X-100, 0.5% sodium deoxycholate, 0.1% SDS) supplemented with EDTA-free protease inhibitors (cOmplet, Roche) for 30 min, then cells debris were removed by high speed centrifugation at 4°C for 25 min. Protein concentration was measured by spectrophotometry using Bio-Rad Bradford protein assay reagent. Samples were denatured at 95°C for 10 min in Laemmli buffer (Bio-Rad) with 2% β-mercaptoethanol. Samples were subjected to SDS-PAGE for Western blot analysis.

To asses RBMX protein levels after overexpression in HEK293, cells were seeded at a density of 62,500 cells/cm² per well of a P6-well plate and transfected using jetOPTIMUS® DNA Transfection Reagent (Polyplus) with a 1:1 ratio of DNA/polymer according to the manufacturer’s protocol. Cells were transfected with 0.5 µg of pCAGEN empty vector (negative control) or a pCAGEN vector containing the CDS for human wildtype or patient variant RBMX or RBMXL1 with an N-terminal HA-tag. 24 hours post-transfection, cells were incubated for 5 minutes at room temperature in NP-40 buffer (500 mM NaCl, 50 mM HEPES, 1% NP-40) for lysis. Cell debris was removed by centrifugation for 5 minutes at 4°C at 12,000 x g. The protein amount in the supernatant was measured with Pierce™ BCA Protein Assay Kit (Thermo Scientific). 7.5 µg protein were mixed with 4x Laemmli (Bio-Rad) with 2% 2-mercaptoethanol (Sigma-Aldrich), denatured for 10 min at 95°C and subjected to SDS-PAGE for Western blot analysis.

### Immunoprecipitation assay

For targeted mass spectrometry (PRM), hNSCs were seeded at a density of 15,000 cells/cm² in Petri dishes. To induce neuronal differentiation, hNSCs were initially maintained for 2 days in complete medium containing BDNF, EGF, FGF-2, and gentamicin. After two days, the medium was replaced with differentiation medium composed of the same base medium supplemented only with BDNF and gentamicin. Cells were maintained under these differentiation conditions for 19 days, with medium changes every two days. For immunoprecipitation (IP), hNSCs and neurons were rinsed once with cold 1× PBS on ice. Cells were lysed in 400 µL of lysis buffer [50 mM Tris·HCl, pH 7.4, 100 mM KCl, 1 mM MgCl_2_, 0.1 mM CaCl_2_, 0.1% SDS, 1% NP40, 0.5% sodium deoxycholate, 1x cOmplete EDTA-free protease inhibitor cocktail and 30 U of RNase inhibitors (RNAsin, Promega)]. Lysates were centrifuged for 15 min at 13,000 rpm at 4°C, and the supernatants were pre-cleared by sequential incubation with 25 μL of free Dynabeads protein G (ThermoFisher Scientific), followed by 25 μL of Dynabeads protein G coupled to 2.5 μg of rabbit anti-mouse IgGs (Jackson ImmunoResearch, 315-005-044), each for 1 hour. The pre-cleared lysates were then incubated overnight at 4°C with agitation with 25 μL of Dynabeads protein G coupled to 2.5 μg of anti-*RBMX/RBMXL1* antibody (ab190352, Abcam). After IP, the beads were washed five times with 500 µL of high-salt wash buffer [50 mM Tris·HCl, pH 7.4, 1 M KCl, 1 mM EDTA, 1% NP40, 0.5% sodium deoxycholate, and 0.1% SDS; 4°C], followed by five washes with 1X PBS. Beads were dried and stored at −20°C until proteomic analysis.

For IP experiments performed to identify RBMX/RBMXL1 partners, as well as co-IP experiments conducted to validate the mass spectrometry data, HEK293T cells at 50-60% confluency were transfected with 2.5 μg of plasmid (HA-tagged WT RBMX, WT RBMXL1, empty vector (pCAG) or HA-p.R381* for the co-IP experiments) using Lipofectamine 2000 (ThermoFisher Scientific), according to the manufacturer’s protocol. After, 24 hours post-transfection, cells were rinsed once with cold 1× PBS on ice. HEK293T from 4 wells of 6-well plate were lysed in 800 µL of lysis buffer [50 mM Tris·HCl, pH 7.4, 100 mM KCl, 1 mM MgCl_2_, 0.1 mM CaCl_2_, 0.1% SDS, 1% NP40, 0.5% sodium deoxycholate and 1x cOmplete EDTA-free protease inhibitor cocktail]. Samples prepared for mass spectrometry analysis were not treated with RNase inhibitors or RNase, whereas co-IP samples were treated with either 30 U of RNase inhibitors (RNAsin, Promega) or with 8 μl of RNase A (10 mg/ml) (ThermoFisher Scientific), added in the lysis buffer. Lysates were centrifuged for 15 min at 13,000 rpm at 4°C, and supernatants were pre-cleared by sequential incubation with 50 μL of free Dynabeads protein G (ThermoFisher Scientific), followed by 50 μL of Dynabeads protein G coupled to 2.5 μg mouse anti-rabbit IgGs (Jackson

ImmunoResearch, 211-002-171), each 1 hour. The pre-cleared lysates were then incubated overnight at 4°C with agitation with 25 μL of anti-HA Magnetic beads (88836, Thermo Scientific). After IP, the supernatants were saved as ‘inputs’ (HA-RBMX, HA-RBMXL1, empty vector (pCAG) or HA-p.R381*), and the beads were washed five times with 500 µL of high-salt wash buffer [50 mM Tris·HCl, pH 7.4, 1 M KCl, 1 mM EDTA, 1% NP40, 0.5% sodium deoxycholate, and 0.1% SDS; 4°C]. Beads intended for mass spectrometry analysis were washed five times with 1X PBS, dried and stored at −20°C until proteomic analysis. The beads used for co-IP experiments were resuspended in 50 μL of loading buffer [120 mM Tris⋅HCl, pH 6.8, 8% SDS, 31% glycerol, 1.4 M β-mercaptoethanol, and bromophenol blue] and incubated for 5 min at 95°C. Co-IP samples (HA-RBMX, HA-RBMXL1, empty vector (pCAG), or HA-p.R381*) treated with RNAsin or RNAse A were subjected to SDS-PAGE for Western blot analysis. For Co-IP experiments performed to asses RBMX variant/RBMXL1 binding capacity, HEK293 cells were seeded at a density of 62,500 cells/cm^2^ per well of a P6-well plate. At 50-60% confluence, three wells per condition were transfected with a total of 1.25 μg pCAGEN empty vector + 1.25 µg HA-RBMX (negative control) or 1.25 µg FLAG-RBMXL1 + 1.25 µg HA-RBMX variants per condition. Transfection was carried out with jetOPTIMUS® DNA Transfection Reagent (Polyplus) with a 1:1 ratio of DNA/polymer according to the manufacturer’s protocol. 24 hours post transfection, Co-IP was performed with the µMACS™ DYKDDDDK Isolation Kit and µColumns (both Miltenyi Biotec) according to the manufacturer’s instructions with minor modifications. To prevent unspecific binding of HA-RBMX to the paramagnetic beads, 50 µl of anti-DYKDDDDK beads were blocked with 50 µl 5% BSA (Roth) in 1x TBS (Thermo Fisher) with 0.1% Tween 20 (Sigma-Aldrich) for 1 hour at 4°C with rotation. Cell pellets were lysed in 400 µl Lysis buffer [150 mM NaCl, 1% Ecosurf™ EH-9, 50 mM Tris HCl (pH 8.0)] with 1x Halt Protease Inhibitor Cocktail (Thermo Scientific) for 30 minutes, cell debris was removed by high-speed centrifugation for 10 minutes at 4°C and 10,000 x g. 60 µl of supernatant were mixed with 4x Laemmli (Bio-Rad) with 2-mercaptoethanol (Sigma-Aldrich) and labelled as ‘input’. The remaining supernatant was mixed with 100 µl of blocked anti-DYKDDDDK beads/5% BSA TBST mixture and incubated for 30 min at 4°C with rotation. µColumns were pre-equilibrated with 200 µl of Lysis buffer with 1x Halt Protease Inhibitor, samples were transferred onto µColumns and washed 4x with high-salt Wash Buffer 1 [500 mM NaCl, 1% Ecosurf EH-9, 0.5% sodium deoxycholate, 0.1% SDS, 50 mM Tris HCl (pH 8.0)] with 1x Halt Protease Inhibitor and once with 100 µl of Wash Buffer 2 [20 mM Tris HCl (pH 7.5)]. Proteins were eluted by adding 20 µl of 95°C hot Elution Buffer [50 mM Tris HCl (pH 6.8), 50 mM DTT, 1% SDS, 1 mM EDTA, 0.005% bromphenol blue, 10% glycerol] to columns, incubation for 5 minutes at room temperature and subsequent elution by addition of another 50 µl of hot Elution buffer. Samples were then labelled ‘Co-IP’ and together with input samples denatured for 10 min at 95°C. Samples were stored at −20°C until subjected to SDS-PAGE for Western blot analysis.

### Western blot

Samples were resolved by SDS–PAGE and transferred onto PVDF membranes (Merck, #IPVH00010). Membranes were blocked in 5% milk in TBS buffer with 0.1% Tween (TBS-T) and incubated overnight at 4°C with the appropriate primary antibody in blocking solution. Membranes were washed three times in TBS-T, incubated at room temperature for one hour with HRP-coupled secondary antibodies at 1:10,000 dilution in TBS-T, followed by three times TBS-T washes. Visualization was performed by quantitative chemiluminescence using SuperSignal West Pico or Femto PLUS Chemiluminescent Substrate (Sigma).

For quantification of RBMX/RBMXL1 in non-transfected cells (fibroblast lines), HA-RBMX WT and variants and HA-RBMXL1 in transfected cells (HEK293), and FLAG-RBMXL1 binding to HA-RBMX WT and variants after Co-IP experiments (HEK293), samples were resolved by SDS-PAGE. Samples were separated using 8–16% Mini-PROTEAN® TGX™ Precast Protein Gels and transferred onto Trans-Blot Turbo Mini 0.2 µm Nitrocellulose membranes using the Trans-Blot Turbo Transfer System (all Bio-Rad). Membranes were blocked with EveryBlot Blocking Buffer (Bio-Rad) for 10 min with shaking at room temperature and incubated with primary antibodies diluted in 5% BSA (α-Actinin, RBMX) in TBST or 5% milk in PBS (HA, FLAG) at 4°C overnight with shaking. Membranes were washed five times for 10 min in TBS-T, incubated at room temperature for one hour with HRP-coupled secondary antibodies at 1:50,000 dilution in 5% BSA in TBS-T, followed by five 10-minute TBS-T washes. Visualization was performed by quantitative chemiluminescence using SuperSignal West Dura Extended Duration Substrat (Thermo Scientific). Imaging was done with the ChemoStar Touch (Intas).

Signal intensity was quantified using ImageJ. Primary and secondary coupled HRP antibodies used for Western blot are described in **Supplementary Table 15**. All immunoblot experiments consisted of at least three independent replicates.

### Mass spectrometry

#### Analysis of binding partners

After IP, the proteins retained on the beads were reduced 30 min at 56°C with 5 mM TCEP in Urea 2M (Sigma Aldrich), 0.1M Tris pH8, then alkylated 30 min at RT with 10 mM IAA (iodoacetamide). Trypsin digestion was carried out at 37°C, OVN with 500 ng enzymes. After acidification with Trifluoroacetic acid (TFA) (Sigma Aldrich), the peptide supernatant was desalted on C18 spin column and dried on Speed-Vacuum before LC-MS/MS analysis. Samples were analyzed using an Ultimate 3000 nano-RSLC (Thermo Scientific, San Jose California) coupled in line with a quadrupole - orbitrap Exploris 480 via a nano-electrospray ionization source (Thermo Scientific, San Jose California) and the FAIMS pro interface. The tryptic peptides were loaded on a C18 Acclaim PepMap100 pre-column (75 µm x 2 cm, 3 µm, 100Å, Thermo Fisher Scientific) for 1 minute at 15 µL/min with 2% Acetonitrile MS grade (ACN) (Sigma Aldrich), 0.1% Formic acid (FA) in H_2_O and then separated on a C18 PepMap nano-column (75 µm ID x 25 cm, 2.6 µm, 150Å, Thermo Fisher Scientific) with a 23 minutes linear gradient from 7% to 35% buffer B (A: 0.1% FA, in H_2_O; B: 0.1% FA in 80% ACN) at 400 nL/min followed by a regeneration step at 90% B and a equilibration at 7% B. The total chromatography was 30 minutes. The mass spectrometer was operated in positive ionization mode in Data-Dependent Acquisition (DDA) for one FAIMS compensation voltages (CV = −45V). The DDA cycle consisted of one survey scans or MS1 (300-1500 m/z, 120,000 FWHM) followed by MS² spectra (HCD; 30% normalized energy; 2 m/z window; 30,000 FWMH) in the limit of 1.2 sec. The Automatic Gain Control (AGC target) were set to 3E6 and 1E5 respectively for MS1 and MS2 scans, and the maximum injection time (IT) was set to 50 ms for both scan modes. Unassigned and single charged states were rejected. The exclusion duration was set for 40 s with mass width was ± 10 ppm. Proteins were identified with Proteome Discoverer 2.5 software (Thermo Fisher Scientific) and Homo Sapiens proteome database (UniProt, reviewed, release 2024_04_03 with 20598 entries). Precursor and fragment mass tolerances were set at 10 ppm and 0.02 Da respectively, and up to 2 missed cleavages were allowed. Oxidation (M) was set as variable modification, and Carbamidomethylation (C) as fixed modification. Peptides were filtered with a false discovery rate (FDR) at 1%, rank 1. Proteins were quantified with a minimum of 1 unique peptide based on the XIC (sum of the Extracted Ion Chromatogram). The quantification values were exported in Perseus for statistical analysis involving a log[2] transform, imputation, normalization^51^. Gene Ontology (GO) analysis was performed using DAVID tool^52^ on the top proteins (*P*-value<0.05) identified by mass spectrometry data. The top 10 categories were selected based on Benjamini– Hochberg-corrected *P*-values. A correlation plot was generated by calculating the difference in binding (log₂ fold change) between HA-RBMXL1 and HA-RBMX conditions for each shared interactor (*P*-value<0.05) identified in the mass spectrometry data. Each point represents a shared interactor, plotted according to its log₂ fold change in binding to HA-RBMX (x-axis) and HA-RBMXL1 (y-axis). A linear regression was fitted to the data, and the coefficient of determination (*R*²) was calculated to assess the strength of the correlation between the two conditions.

#### Analysis of human RBMX and RBMXL1 relative protein abundance

The same analysis than the one for binding partners was performed, except that it was used with an inclusion mass list. The mass list contains (compound: GLPPSMER, m/z: 443.725; IVELLMK, 472.796; IVEVLLM(-Ox)K, 480.793; VEQATKPSFESGR, 718.362; GLPPSVER, 427.739; IVEVLLIK, 463.817; VEQATKPSFER, 646.335). If no targets of the previous list were found, dependent scan was performed on the most intense ion. For RBMX, all the compounds can be used to detect both RBMX isoforms (NP_002130.2 and NP_001158275.1), but only the following compounds, IVELLMK and IVEVLLM(Ox)K, are found to the isoform NP_001158275.1.

### Image acquisition and analysis of *in utero* electroporation experiments

Images for *in utero* electroporation experiments were acquired using a TCS SP8 X (Leica microsystems) confocal microscope using a 20x DRY HC PL APO CS2 objective. For all experiments, a Z-stack of 1 μm was acquired and the image size was 1024x1024. In order to analyse our images, we use FIJI software^53^ to select the area of interest (electroporated region with GFP-positives cells), the images are saved and a project is created in QuPath.^54^ Cellpose^55^ in QuPath is used to detect the GFP-positives electroporated cells). We then exported a CSV file containing the number of GFP-positive electroporated cells in each cortical area to calculate the percentage of positive cells for each image. Cell counting was carried out on two to four different brain sections from at least three different embryos per condition. Only male embryos were analyzed. Only similarly electroporated regions were included in the analysis. Cortical areas (cortical plate (CP), intermediate zone (IZ), and subventricular/ventricular zones (SVZ/VZ)) were delineated based on cell density (nuclei count with DAPI staining). All experiments were performed in at least three independent biological replicates.

### Statistics

All statistics analyses were performed using GraphPad Prism 6 (GraphPad) or R and are represented as mean +/- S.E.M for bar graph. Box plot elements are defined as follows: center line: median; box limits: upper and lower quartiles; whiskers: 1.5× interquartile range; points: outliers. The level of significance was set at *P* < 0.05 in all the statistical tests. All statistical tests used and n size numbers are shown in the figure legends and statistical details are reported in **Supplementary Table 16**. Analysis of IUE experiments and neuroanatomical characterization of mouse brains were performed blinded. We applied parametric tests, which assume normality and homogeneity of variances. The normality assumption was evaluated and supported by Quantile-Quantile (QQ) plots (**Supplementary Table 16**).

### Data availability

Variant details will be submitted to ClinVar for publication. RNA-seq will be deposited in the European Genome-phenome Archive (EGA, http://www.ebi.ac.uk/ega), hosted by the EBI. Both datasets will be subject to a Data Processing Agreement, and access requests will be reviewed by a Data Access Committee to ensure compliance with ethical and legal standards. The mass spectrometry proteomics data have been deposited to the ProteomeXchange Consortium via the PRIDE partner repository with the dataset identifier PXD066823. High-throughput sequencing data performed in N2A cells has been deposited in the Gene Expression Omnibus Database (GSE305608). All other data are available in the manuscript or its supplementary files.

### Code availability

No custom code was developed for this study.

## Acknowledgments

We thank the patients and their families for their participation in this study. Patients included in this study were diagnosed through projects supported by Agence National de la rechercher (ANR)-project ANR-13-BSV1-0027 (CILAXCAL) to T.A-B (and CD as partner). Patient 8 was identified through the DECIPHER database (https://deciphergenomics.org, patient ID: 488393). This project was financially supported by Deutsche Forschungsgemeinschaft (DFG) -Project number 505514143 to CD, Agence National de la rechercher (ANR)-Project RETRO-RBMX (ANR-22-CE92-0072) to JDG, the program Investissements d’Avenir labeled (ANR-10-IDEX-0002-02, ANR-10-LABX-0030-INRT, to JDG), SFRI-STRAT’US project [ANR 20-SFRI-0012] to JDG, INSERM/CNRS and University of Strasbourg to JDG, Förderverein Universitätsneurologie e.V. and Stiftung Universitätsmedizin Essen. P. Tilliole is supported by Ministère de l’Enseignement Supérieur de la Recherche et de l’Innovation and Fondation pour la recherche médicale. E.E.D. is the Ann Marie and Francis Klocke, MD Research Scholar. P.Tilly is research assistant at the University of Strasbourg. J.D.G. is an INSERM investigator. N.S and J.F.T are supported by ERC CoG 2021 TransNeuroFate (to JDG). J.K.C. was supported by a Fonds de Recherche du Québec - Santé (FRQS) graduate fellowship, W.A.P. was supported by an FRQS Chercheur-boursier award. S.C.C. is a Senior Lecturer at the University Bourgogne Europe. B.Y. is an INSERM investigator supported by grants from the Agence Nationale de la Recherche and the IFCPAR/CEFIPRA (grant no. 6503-J). We thank the Imaging Center Essen (IMCES) at the Faculty of Medicine of the University of Duisburg-Essen, Germany for providing access to Zeiss Elyra PS.1 and especially Dr. Anthony Squire for general support in its operation. We acknowledge the Imaging Center of IGBMC (https://ici.igbmc.fr/) part of the IBiSA labeled PIQ-QuESt (https://piq.unistra.fr) and member of the national infrastructure France-Bioimaging (https://france-bioimaging.org) supported by the French National Research Agency (ANR-10INBS-04). We are grateful to the staff of the molecular biology service (in particular, Thierry Lerouge and Paola Rossolillo), of the mouse facilities of the Institut Clinique de la souris (ICS) (in particular Sophie Brignon), of the GenomeEast platform (part of France Genomic infrastructure), of the flow cytometry service and of the IGBMC PluriCell East platform (in particular Amélie Freismuth and Charlotte Fugier-Schmucker) for their involvement in the project. We also thank Doulaye Dembele from the IGBMC GenomEast sequencing platform for his help with statistical analysis. We are also grateful to members of J.D.G.’s and C.D’s laboratories for discussion and technical assistance. We warmly thank Dr. Martin Grosse (Institute for Human genetics, UK Essen) and Dr. Clement Charenton (IGBMC, Strasbourg, France) for critical discussion, Dr. Christopher Schröder and Dr. Fabian Kilpert (Institute for Human genetics, UK Essen) for bioinformatics analyses of RNAseq data (patients’ fibroblasts), Dr Ting Cai and Stephane Richards (McGill University, Montréal, Canada) for sharing iPSCs with p.R355Kfs*8, Dr. Maria Wilbe, Pr. Marie-Louise Bondeson and Dr. Josefin Johansson (Uppsala University, Uppsala, Sweden) for sharing data and reagents, and Pr A. Firquet and Pr Dr S. Perrier-d’hauterive at the CHC hospital and R. Close at the Nguyen lab for the collection of human brain samples. We are grateful to Valérie E. Vancollie and Christopher J. Leliott, as well as members of the Sanger Institute Mouse Pipelines teams and the Research Support Facility, for the management of the *Rbmx*^tm2a(KOMP)Wtsi^ knockout mouse line used in this study. We thank Dorinda Wright and Stephanie Littlewood for histological work at HistologiX Ltd, and the students and technicians of the Yalcin’s laboratory for their contributions to the neuroanatomical studies, in particular Emeline Aguilar, Christel Wagner, Marie-Christine Fischer, Anna Mikhaleva, Rebecca Balz, and Léo Gagliardi. E.A., E.S., D.H., T.A-B., B.Y. and C.D. are members of the International Research Consortium for the Corpus Callosum and Cerebral Connectivity (IRC^5^) consortium (https://www.irc5.org/). This study makes use of data generated by the DECIPHER community. A full list of centres who contributed to the generation of the data is available from https://deciphergenomics.org/about/stats and via email from. DECIPHER is hosted by EMBL-EBI and funding for the DECIPHER project was provided by the Wellcome Trust [grant number WT223718/Z/21/Z]. Support for title page creation and format was provided by AuthorArranger, a tool developed at the https://authorarranger.nci.nih.gov/.

## AUTHOR CONTRIBUTION

C.D., C.N and A.R. performed the genetic analyses and C.D. collected molecular and clinical data. L.B., B.I.D., D.L., I.A., S.L.T., B.C., P.C., C.E.P., S.J.L., S.A., T.N.K. E.E.D., E.A., W.D., E.S., A.v;H., D.H., V.C-D., T.A-B. contributed molecular and clinical data from other cohorts. E.Leitão performed the evolutionary analysis. P.Tilliole and P.Tilly. conceived, performed and analysed IUE experiments. S.K. and C.M. performed analysis of patients’ samples. T.C., E.Lejeune, and B.K. performed RNAseq experiments in patients. Y.M. performed RNAseq in N2A cells. D.P. analysed RNAseq data (N2A). P.Tilliole and C.M conceived and performed *in vitro* functional assays. N.Schwaller performed Western-Blot and immunohistochemistry, J-F.T. and M.J. performed expression analysis from published datasets. M.D.V. and E.G. built the automatic tool for analysis of IUE. B.M. performed mass spectrometry analysis and analyzed data. L.S. provided control cell lines and helped with functional experiments. M.L. and N.Sestan performed analysis of brainspan data. S.C.C. conducted X-ray image analyses of the tm2a mouse line; and H.W., S.C.C., and B.Y. performed the neuroanatomical characterisation of the tm2a mouse line. M.R. validated the tm2a mouse model. S.T., E.E. and L.N. contributed data from human embryo. J.K.C, and W.A.P helped with IPSCs culture. P. Tilliole, C.M., E. Leitão developed bioinformatics pipelines, performed data analyses and made the final figures. C.D. and J.D.G. conceived and supervised the project and drafted the article. All authors reviewed and approved the final manuscript.

## SUPPLEMENTARY FIGURE LEGENDS

**Supplementary Fig. 1. Brain MRI of Patients. a,** MRI of patient 5 showing severe microcephaly with foreshortened frontal lobes, diffuse dysgyria/microgyri (perisylvian-predominant), extensive periventricular nodular heterotopia, mildly enlarged extra-axial spaces, short dysplastic corpus callosum, reduced white matter and mild cerebellar vermis hypoplasia. **b,** MRI of patient 4. Findings include inferior vermian hypoplasia with widening at the level of the obex. The brainstem appears small in size. The corpus callosum shows a slightly dysgenetic configuration, with thinning of the posterior body and splenium. A simplified gyral pattern is evident across much of the bilateral cerebral hemispheres, with mild interdigitation of the gyri noted in the parasagittal frontal lobes. A persistent cavum septum pellucidum is present. The lateral, third, and fourth ventricles are mildly dilated. **c,** MRI of patient 1 showing ventriculomegaly and a hypotrophy of the white matter. **d,** MRI of patient 2 showing a mild prominence of trigones and occipital horns of the lateral ventricles resulting in a mild colpocephalic configuration.

**Supplementary Fig. 2. Expression of RBMX in patients’ fibroblasts**. **a**, *RBMX* expression in patient fibroblasts versus controls measured by RT-qPCR. *RBMX* expression was normalized to *PPIA* and to the mean of controls per experiment (*n*=3 / three technical replicates each). *P*-values were calculated by Mann-Whitney U test and are indicated above comparisons. **b**, Localization of specific human *RBMX* (upper panels) and *RBMXL1* (lower panels) RT-qPCR primers. Primer specificity was validated by Sanger sequencing. Representative sequencing traces are shown for each primer set. **c**, Immunofluorescence staining of RBMX/RBMXL1 in patient and control fibroblasts. RBMX/RBMXL1 is shown in green (pseudo-colored) and nuclei are stained with DAPI (blue). Images show representative fields (*n*=1 per sample). Scale bar: 10 µm.

**Supplementary Fig. 3. Comparative analysis RBMX and its retrocopies in human and mouse and characteristics of *Rbmx^tm2a^*mouse. a,** Evolutionary distances between human RBMX and RBMXL1 and their mouse counterparts Rbmx, Rbmxl1a, and Rbmxl1b. Scale bar depicts the number of substitutions per site. **b,** Sequence alignments of human RBMX and RBMXL1 alongside mouse Rbmx, Rbmxl1a, and Rbmxl1b, with differences highlighted using colors.

**Supplementary Fig. 4. Validation of *Rbmx^tm2a^* mice and *Rbmx* expression using LacZ reporter. a**, Localization of specific mouse *Rbmx* (upper panels) and *Rbmxl1* (lower panels) RT-qPCR primers. Primer specificity was validated by Sanger sequencing. Representative sequencing traces are shown for each primer set. **b,** Quantification by RT-qPCR of *Rbmx* (left panel) and *Rbmxl1* (right panel) mRNA levels in cortices isolated from E15.5 *Rbmx^tm2a^* males. Gene expression was normalized to *Rplp0* and to the mean of WT per experiment (*n*=6 per condition) and analysed using Unpaired *t*-test. *P*-values are indicated above for comparison. **c, d,** Phenotypic characterization of the *Rbmx^tm2a^* mouse. **c,** Expression analysis in adult males (16 weeks old) using the lacZ reporter cassette. Staining was performed in three independent male samples. **d,** Histopathological examination of the eye in six independent adult *Rbmx*^tm2a^ male mice aged 15–16 weeks. Scale bar, 500 µm. **e**, Description of cephalometric measurements performed in *Rbmx^tm2a^* mice.

**Supplementary Fig. 5. *RBMX* and *RBMXL1* have similar temporal and spatial expression patterns in human brains. a**, **b**, Expression pattern of *RBMX* or *RBMXL1* in human across (**a**) brain regions or (**b**) selected cell types of the cerebral cortex (x axis) and ages (y axis). Data obtained by multiple published datasets through https://cellxgene.cziscience.com.^18^ Color scale represents the weighted average expression across datasets and dot size represents the weighted average percentage of cells expressing *RBMX* (red) or *RBMXL1* (blue). **c**, Analysis of available single-cell RNA sequencing data from gestational week (GW) 16 human cortices^19^ showing the percentage of neurons expressing *RBMX* and *RBMXL1* individually or simultaneously. Of note, 0.4% of neurons express neither gene. **d**, Quantification by Parallel Reaction Monitoring (PRM) of the relative expression of RBMX and RBMXL1 proteins in human neural stem cells (hNSCs) and neurons derived from hNSCs. XIC (extracted ion chromatogram) values are used to quantify protein abundance. Each point represents data from an independent culture. Data (mean ± SEM) were analyzed using Unpaired *t*-test. *P*-value is indicated above for comparison. **e**, Schematic representation of the location of the specific peptides used in PRM to determine the RBMX/RBMXL1 ratio shown in **d**.

**Supplementary Fig. 6. RBMX and RBMXL1 shared protein and mRNA interactors. a**, Immunostaining for HA (red), GFP (green) in human neural stem cells (hNSCs) after transfection with wildtype (wt) HA-RBMX or HA-RBMXL1 and pCAG-GFP constructs show nuclear localization of both proteins. Cells were counterstained with DAPI (blue). Scale bars: 30 μm, insets: 5 μm. **b**, Western blot analysis of HA immunoprecipitation (IP) experiments performed after incubation of an RNase inhibitor (RNasin, upper panel) or RNase A (lower panel) in HEK293T cells transiently transfected with the indicated constructs. Whole-cell extracts are shown as input. **c**, Quantification of the ratios of co-immunoprecipitated proteins relative to the amounts of HA-tagged proteins from 3 independent experiments indicate a similar binding of HA-RBMX (blue) and HA-RBMXL1 (orange) to endogenous MATR3, HNRNPDL, TRA2B, and SRSF7 proteins. Data (mean ± SEM), normalized to the mean of HA-RBMX condition, were analyzed using Unpaired *t*-test. *P*-value is indicated above for comparison. **d**, CLIP performed in hNSCs overexpressing HA-RBMX (blue) and HA-RBMXL1 (orange) followed by qPCR for candidate genes show enrichment of *ASPM*, *ATRX*, *SMN1/2* and *HP1α* mRNA but not of *18S ribosomal RNA* used as a control. Bar charts represent IP/Input enrichment relative to the control condition (empty vector) from 3 biological replicates. Data (mean ± SEM) were analyzed using one-way ANOVA, with Bonferroni’s multiple comparisons test. Adjusted *P*-values are indicated above for comparison (to empty vector condition).

**Supplementary Fig. 7. Strategies for *SMN2* minigene and *ATRX* exitron assays. a,** Schematic representation of alternative splicing of *SMN2* exon 7 regulated by RBMX. The top panel shows the *SMN2* minigene construct containing exons (Ex) 6, 7, and 8 with full-length introns in-between. In the presence of RBMX (+RBMX), exon 7 is included during splicing, generating mRNA with exons 6, 7, and 8 (*SMN2*). In the absence of RBMX (-RBMX), exon 7 is skipped, generating mRNA comprising only exons 6 and 8 *(SMN2Δex7*). **b**, **c**, Western blot (**b**) and quantification (**c**) of HA-RBMX and HA-RBMXL1 protein after overexpression in HEK293 cells. **b,** α-Actinin served as a loading control; molecular masses in kilodaltons (kDa). **c**, Boxplots show the ratio of HA-RBMX to α-Actinin signal intensity normalized to the mean of HA-RBMX samples (*n*=3). Statistical significance was assessed using Unpaired t-test. *P*-value is shown above for comparison. **d**, Schematic of the workflow for *in-vitro ATRX* exon 9 splicing assay in HEK293. Cells are seeded on day 1 and transfected on day 2 with either control siRNA or siRNA targeting the 3′UTR of *RBMX* to induce *ATRX* exon 9 exitron formation. On day 3, cells are transfected with either empty vector (negative control), HA-tagged RBMX, or HA-tagged RBMXL1 for rescue. On day 4, *ATRX* exitron inclusion is assessed by (**e**) RT-PCR and quantified by (**f**) RT-qPCR. Detection (**e**) and quantification (**f**) of *ATRX* exon 9 exitron inclusion in HEK293 cells after endogenous *RBMX* knockdown and transfection with an empty vector. (**e**) Upper panel: RT-PCR strategy using primers in exon 9 to detect all *ATRX* isoforms and primers in exitron to detect full length isoform. Lower panel: Agarose gel showing PCR products for *ATRX* with exitron inclusion (*ATRX* full-length exon 9) and with exitron exclusion (*ATRX* short exon 9). *HPRT1* is used as a cDNA control. (**f**) Upper panel: RT-qPCR strategy using primers flanking the exitron in exon 9 to detect both short and full length *ATRX* isoforms. Lower panel: Boxplots of *ATRX* full-length exon 9 normalized to total *ATRX* exon 9 amounts based on RT-qPCR data after treatment of HEK293 cells with non-targeting siRNA (siRNA Control) or siRNA targeting *RBMX* 3’UTR (*n*=3). Data are normalized to the mean of siRNA Control samples. Statistical significance was assessed using Unpaired *t*-test. *P*-value is shown above for comparison. **g**, **h**, RBMX knockdown in HEK293 cells reduces *RBMX* expression without affecting *RBMXL1*. **g,** Boxplots of *RBMX* expression after treatment of HEK293 cells with non-targeting siRNA (siRNA Control) or siRNA targeting *RBMX* 3’UTR based on RT-qPCR data (*n*=3). Data are normalized to the mean of siRNA Control samples. Statistical significance was assessed using Unpaired *t*-test. *P*-value is shown above for comparison. **h,** Boxplots of *RBMXL1* expression after treatment of HEK293 cells with non-targeting siRNA (siRNA Control) or siRNA targeting *RBMX* 3’UTR based on RT-qPCR data (*n*=3). Data are normalized to the mean of siRNA Control samples. Statistical significance was assessed using Unpaired *t*-test. *P*-value is shown above for comparison.

**Supplementary Fig. 8. *Rbmx* and *Rbmxl1* expression pattern overlap in the mouse developing cortex. a**, E13.5 and E15.5 embryonic mouse brain coronal and sagittal sections immunolabeled for Rbmx/Rbmxl1. Scale bars: 200 μm. Digital magnifications represent Rbmx/Rbmxl1 expression in the cortex: VZ: ventricular zone, IZ: intermediate zone, CP: cortical plate. **b**, Representative Western blot showing Rbmx/Rbmxl1 expression in extracts from cortices of CD1 WT mice from E12.5 to P21. Gapdh immunolabelling was used as loading control. **c**, Quantification of Rbmx/Rbmxl1 expression levels normalized to Gapdh protein levels. AU: Arbitrary Unit. Data (mean ± SEM) from 5 independent experiments (*n*=5 animals per developmental stage) were analyzed using two-way ANOVA, with Bonferroni’s multiple comparisons test. ns: non-significant. Adjusted *P*-value is indicated above for comparison. **d**, Quantification by RT-qPCR of *Rbmx* (left panel) and *Rbmxl1* (right panel) mRNA in male mouse cortices at different stages (*n*=6). AU: Arbitrary Unit. Data (mean ± SEM) from 6 animals per developmental stages were analyzed two-way ANOVA, with Bonferroni’s multiple comparisons test, ns: non-significant. **e**, Analysis of available single-cell RNA sequencing data^23^ from mouse cortices showing the percentage of E13 apical progenitors and neurons (born at E13) expressing *Rbmx* and *Rbmxl1* individually or simultaneously.

**Supplementary Fig. 9. miRNA-mediated knockdown specifically reduces RBMX or RBMXL1 levels without affecting baseline cortical distribution of electroporated cells. a,** Quantification by RT-qPCR of *Rbmx* (left panel) and *Rbmxl1* (right panel) mRNA levels in Neuro-2a cells transiently transfected with pCAG:miRNA scramble or a pool of pCAG:miRNAs targeting mouse *Rbmx* (#1, #2, #3) or *Rbmxl1* (#1, #2) showing the specificity of the miRNA strategy. The relative expression of *Rbmx* and *Rbmxl1* is normalized to *Rplp0*. AU: Arbitrary Unit. Data (mean ± SEM) from 3 independent experiments were normalized to the mean of controls (miRNA scramble) and analyzed using one-way ANOVA, with Bonferroni’s multiple comparisons test. Adjusted *P*-values are indicated above for comparison (to miRNA scramble). **b**, RNA sequencing analysis of GFP+ Neuro-2a cells transiently transfected with pCAG:GFP and pCAG:miRNAs targeting mouse *Rbmxl1* (*Rbmxl1* #1) or *Rbmx* (*Rbmx* #1). MA-plot shows the log_2_ mean normalized counts of all genes against their log_2_ fold change (*Rbmxl1*-KD versus *Rbmx*-KD (knockdown)). Significant genes identified using an adjusted *P*-value threshold of < 0.05 are represented by red dots. Data from 4 independent experiments were analyzed using Wald test with *P*- values adjusted for multiple testing using the Benjamini and Hochberg method. **c**, Quantification by RT-qPCR of *Rbmx* (left panel) and *Rbmxl1* (right panel) mRNA levels in Neuro-2a cells transiently transfected with pCAG:miRNA scramble or with pCAG:miRNAs targeting mouse *Rbmx* and *Rbmxl1*, either individually (*Rbmx* #1 or #2; *Rbmxl1* #1 or #2) or simultaneously (*Rbmx+Rbmxl1* #1 or #2) showing the specificity of the miRNAs used in the study. The relative expression of *Rbmx* and *Rbmxl1* is normalized to *Actb*. AU: Arbitrary Unit. Data (mean ± SEM) from 3 independent experiments normalized to the mean of control (miRNA scramble) and analyzed using one-way ANOVA, with Bonferroni’s multiple comparisons test. Adjusted *P*-values are indicated above for comparison (to miRNA scramble). **d**, Coronal sections of E15.5 mouse cortices, two days after *in utero* electroporation (IUE) with pCAG:H2B-eGFP and pCAG:miRNA scramble together with empty vector or human WT RBMX or human WT RBMXL1. GFP-positive electroporated cells are depicted in green. Nuclei are stained with DAPI (blue). Scale bar: 50 μm. **e**, Analysis of the percentage (means ± SEM) of electroporated cells in ventricular/subventricular zone (VZ/SVZ), intermediate zone (IZ), and cortical plate (CP) showing no effect of the overexpression of the constructs in control conditions. Data were analyzed by Two-way ANOVA, with Tukey’s multiple comparisons test. Number of embryos analyzed: miRNA scramble + empty vector, *n*=5; miRNA scramble + RBMX, *n*=5; miRNA scramble + RBMXL1, *n*=4. ns: non-significant.

**Supplementary Fig. 10. Overexpression of the RBMX variants does not show an effect under control conditions. a**, Coronal sections of E15.5 mouse cortices, two days after *in utero* electroporation (IUE) with pCAG:H2B-eGFP and pCAG:miRNA scramble together with pCAG-empty vector or various variants as indicated. GFP-positive electroporated cells are depicted in green. Nuclei are stained with DAPI (blue). Scale bar: 50 μm. **b**, Analysis of the percentage (means ± SEM) of electroporated cells in ventricular/subventricular zone (VZ/SVZ), intermediate zone (IZ), and cortical plate (CP) showing no effect of the overexpression of the RBMX variants in control conditions. Data were analyzed by Two-way ANOVA, with Tukey’s multiple comparisons test. Number of embryos analyzed: miRNA scramble + empty vector, *n*=5; miRNA scramble + p.Q83del, *n*=5; miRNA scramble + p. P162del, *n*=4; miRNA scramble + p.Q345*, *n*=9; miRNA scramble + p.E346Gfs*9, *n*=6; miRNA scramble + p.R355Kfs*8, *n*=5; miRNA scramble + p.R381*, *n*=4. ns: non-significant. **c**, Immunostaining for HA (red), GFP (green) in human neural stem cells (hNSCs) after transfection with wild-type (wt) or mutant HA-RBMX and pCAG-GFP constructs show nuclear localization of all mutated proteins except p.P162del. Cells were counterstained with DAPI (blue). Scale bars: 30 μm, insets: 5 μm.

**Supplementary Fig. 11. RBMX variants retain their ability to interact with RBMXL1 and with spliceosomal components. a**, **b**, Western blot (**a**) and quantification (**b**) of HA-RBMX protein after co-immunoprecipitation (Co-IP) of FLAG-RBMXL1 for wildtype and indicated variants. Boxplots show the ratio of HA-RBMX to FLAG-RBMXL1 signal intensity, normalized to the median of wildtype samples (*n*=4). Statistical significance was assessed by Mann-Whitney U test. *P*-values are shown above comparisons. **c**, **d**, Western blot (**c**) and quantification (**d**) of HA-RBMX wildtype and variant protein after overexpression in HEK293 cells. **c**, α-Actinin served as a loading control; molecular masses in kilodaltons (kDa). **d**, Boxplots show the ratio of HA-RBMX to α-Actinin signal intensity normalized on mean of RBMX samples (*n*=3). Statistical significance was assessed using *t*-test and adjusted with Holm correction. Adjusted *P*-values are shown above for comparisons. **e**, Western blot analysis of HA immunoprecipitation (IP) experiments performed after incubation of an RNase inhibitor (RNasin, left panel) or RNase A (right panel) in HEK293T cells transiently transfected with the indicated constructs. Whole-cell extracts are shown as input. **f**, Quantification of the ratios of co-immunoprecipitated proteins relative to the amount of HA-tagged proteins from 3 independent experiments indicate a similar binding of HA-RBMX-p.R381* (green) and HA-RBMX (blue) to endogenous MATR3, HNRNPDL, TRA2B, and SRSF7 proteins. Data (mean ± SEM) were normalized to the mean of HA-RBMX condition and analyzed using Unpaired *t*-test. *P*-values are indicated above for comparison (to HA-RBMX condition). **g**, Quantification by RT-qPCR of *RBMX* (left panel) and *RBMXL1* (right panel) mRNA levels in human neural stem cells (hNSCs) transiently transfected either a control siRNA or a siRNA targeting the ORF of *RBMX* and *RBMXL1* showing the specificity of the siRNA strategy. The relative expression of *RBMX* and *RBMXL1* is normalized to *RPLP0*. AU: Arbitrary Unit. Data (mean ± SEM) from 3 independent experiments were analyzed using Unpaired *t*-test. *P*-values are indicated above for comparison. **h**, Western blot analysis of total RBMX/RBMXL1 protein levels in hNSCs transiently transfected either a control siRNA or a siRNA targeting the ORF of *RBMX* and *RBMXL1* (*n*=2). GAPDH immunolabeling was used as loading control.

